# Unveiling the autoreactome: Proteome-wide immunological fingerprints reveal the promise of plasma cell depleting therapy

**DOI:** 10.1101/2023.12.19.23300188

**Authors:** Aaron Bodansky, David JL Yu, Alysa Rallistan, Muge Kalaycioglu, Jim Boonyaratanakornkit, Damian J. Green, Jordan Gauthier, Cameron J. Turtle, Kelsey Zorn, Brian O’Donovan, Caleigh Mandel-Brehm, James Asaki, Hannah Kortbawi, Andrew F. Kung, Elze Rackaityte, Chung-Yu Wang, Aditi Saxena, Kimberly de Dios, Gianvito Masi, Richard J. Nowak, Kevin C. O’Connor, Hao Li, Valentina E. Diaz, Kaitlin B. Casaletto, Eva Q. Gontrum, Brandon Chan, Joel H. Kramer, Michael R. Wilson, Paul J. Utz, Joshua A. Hill, Shaun W. Jackson, Mark S. Anderson, Joseph L. DeRisi

**Author notes:** These authors contributed equally to this work (David JL Yu, Alysa Rallistan, Muge Kalaycioglu). These authors contributed equally to this work (Paul J. Utz, Joshua A. Hill, Shaun W. Jackson, Mark S. Anderson, Joseph L. DeRisi). Correspondence and requests for materials should be addressed to Mark S. Anderson and Joseph L. DeRisi.

## Abstract

The prevalence and burden of autoimmune and autoantibody mediated disease is increasing worldwide, yet most disease etiologies remain unclear. Despite numerous new targeted immunomodulatory therapies, comprehensive approaches to apply and evaluate the effects of these treatments longitudinally are lacking. Here, we leverage advances in programmable-phage immunoprecipitation (PhIP-Seq) methodology to explore the modulation, or lack thereof, of proteome-wide autoantibody profiles in both health and disease. We demonstrate that each individual, regardless of disease state, possesses a distinct set of autoreactivities constituting a unique immunological fingerprint, or “autoreactome”, that is remarkably stable over years. In addition to uncovering important new biology, the autoreactome can be used to better evaluate the relative effectiveness of various therapies in altering autoantibody repertoires. We find that therapies targeting B-Cell Maturation Antigen (BCMA) profoundly alter an individual’s autoreactome, while anti-CD19 and CD-20 therapies have minimal effects, strongly suggesting a rationale for BCMA or other plasma cell targeted therapies in autoantibody mediated diseases.

## Introduction

Autoantibodies have been identified in a wide range of autoimmune diseases^1–4^. In many cases these autoantibodies are directly pathogenic ^5–10^, while in others they amplify or support T cell driven pathologies^6,11^. Numerous technologies now allow for the detection of autoantibodies to many proteins simultaneously^12–15^, and in the case of phage immunoprecipitation and sequencing (PhIP-Seq), the entire human proteome^16^, using relatively small amounts of plasma or serum. Using these new tools, a wide spectrum of novel autoantibodies have been discovered to be associated with various disease states^11,17–21^, including, but not limited to, paraneoplastic encephalitis, lipodystrophy, inborn genetic disorders, and multisystem inflammatory syndrome in children (MIS-C)^11,16–23^. To accurately identify shared autoreactive profiles among individuals that also share a disease state, we and others have found that serum samples from large numbers of healthy individuals are required, due to the presence of highly diverse autoreactivities present in every individual^17^. However, it remains unclear whether individual autoreactive signatures are stable or variable with time or with immunosuppressive treatment. Here, we leveraged a custom proteome-wide PhIP-Seq autoantibody discovery platform to comprehensively profile the autoreactive repertoire in healthy individuals, and discovered that each individual harbors a unique, distinctive, and highly reproducible set of autoreactivities we term the “autoreactome”. Using longitudinal samples from an additional cohort of healthy individuals, we determined that an individuals’ autoreactome, once formed, remains minimally changed over the course of years.

Extending these findings, we explored the impact of B cell depletion therapies upon the autoreactome. Various surface markers are expressed and then downregulated over the course of B cell development and maturation^24,25^. A subset of these surface markers are targeted by B cell depleting therapies which are used to treat suspected autoantibody mediated diseases, yet a comprehensive evaluation of the effects of these treatments on autoantibodies remains lacking. The most commonly used agent, rituximab, targets CD20, which is not expressed by antibody secreting cells^26^. Recently, chimeric antigen receptor T cell (CAR-T) therapy targeting CD19 positive cells was shown to be safe and effective in the treatment of refractory systemic lupus erythematosus (SLE)^27^. CD19 is expressed on naive and memory B cells, with reduced expression on plasmablasts and plasma cells^28,29^. CAR-T cells targeting B cell maturation antigen (BCMA), which is expressed primarily by antibody secreting plasma cells^30,31^, are approved for the treatment of multiple myeloma^32,33^. Here, we examined the impact of three major B cell depleting therapies, rituximab (anti-CD20), anti-CD19 CAR-T cells, and anti-BCMA CAR-T cells by PhIP-Seq and validate our findings using orthogonal assays. Our findings demonstrate profound impact of anti-BCMA targeted therapies and minimal impact of therapies targeting CD19 or CD20 on individuals’ autoantibody signatures.

### Healthy individuals harbor a unique set of autoreactivities: the “autoreactome”

The search for disease causing or associated autoantibodies is confounded by the fact that autoreactive antibodies are present in all healthy individuals. We have previously shown that large numbers of healthy samples are required to control for this natural confounder and avoid false positive associations^17^. To better understand the variation of the autoreactive antibodies between and within healthy individuals, we obtained serum from 79 pre-COVID healthy blood donors (“Healthy” demographics in Extended Data Table 1) and used our customized, previously described 768,000 element phage immunoprecipitation and sequencing platform(PhIP-Seq)^11,17–21,34^ to determine the proteome-wide set of autoreactivities present within each individual (hereafter referred to as the “autoreactome”).

To determine inter- and intra-individual similarity by PhIP-Seq requires high reproducibility as measured by technical replicate. Over the past several years, we have refined our PhIP-Seq protocols to maximize reproducibility. The master version of this protocol is available at: https://www.protocols.io/view/derisi-lab-phage-immunoprecipitation-sequencing-ph-czw7×7hn?step=14.1. PhIP-Seq protocol performance was evaluated by identifying the PhIP-Seq enrichment similarity of 24 sets of technical duplicates using serum from healthy controls. The raw signal (reads per 100k; rpk) of each phage-presented peptide which was immunoprecipitated within a given sample was calculated, and then used to compare each individual sample. Technical replicates showed high reproducibility (Pearson R-coefficient median =0.946; first quartile (Q1) = 0.907, third quartile (Q3) =0.974) (Fig. 1a: Left Panel).

**Figure 1.**
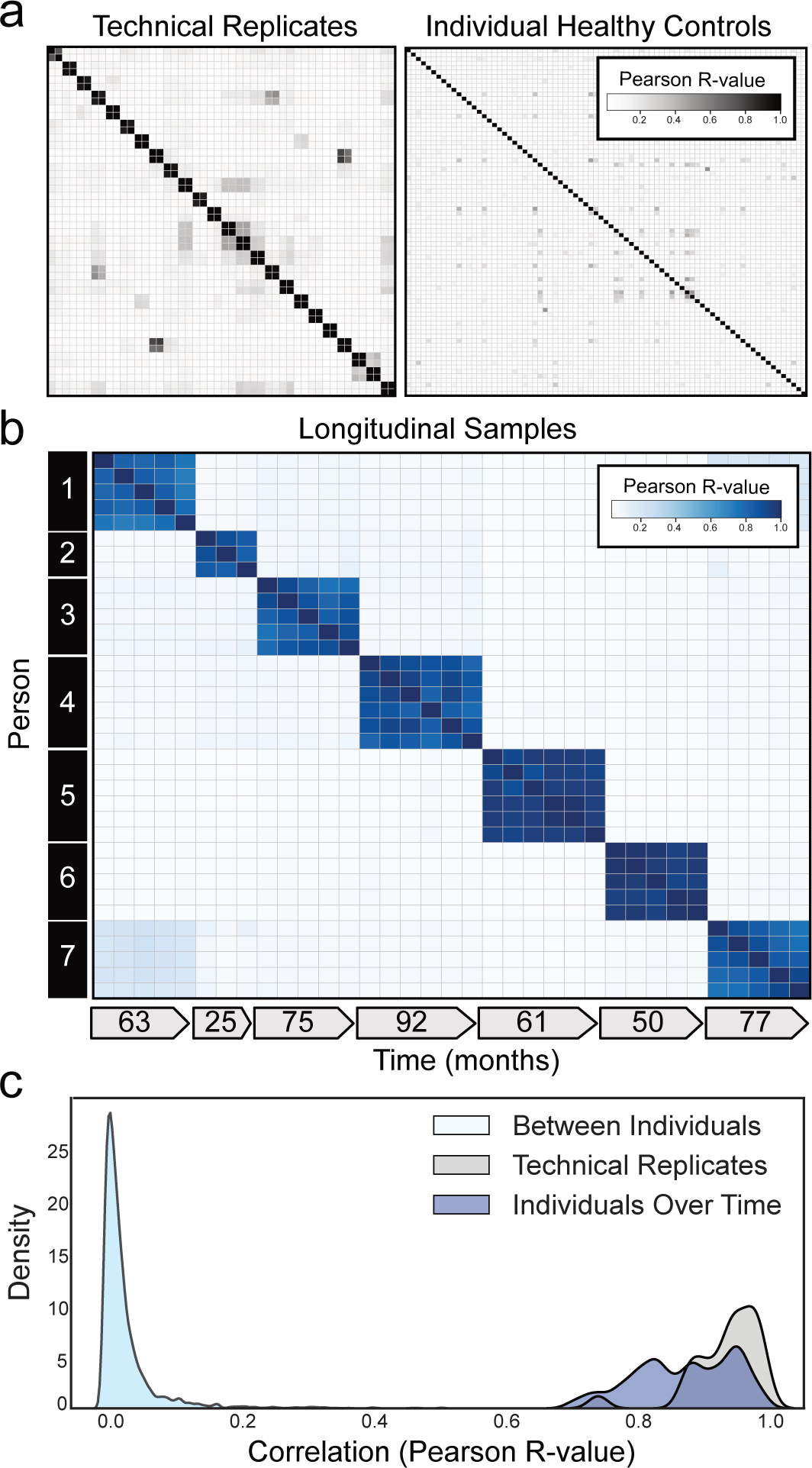
Individuals harbor a unique set of longitudinally stable autoreactivities. **(a)** Correlation matrices showing Pearson correlation coefficients of complete PhIP-Seq signal in healthy individuals. Left: 48 samples representing 24 individuals in technical replicate. Right: 79 distinct individuals. **(b)** Correlation matrix showing Pearson R-values of complete PhIP-Seq signal in 7 distinct individuals each of whom have serial samples over at least 3 years. **(c)** Kernel density estimate plot showing distribution of Pearson R correlation coefficients among technical replicates, individuals over time, and between different individuals.

Next, the similarity of individual autoreactomes was compared between each of the 79 healthy individuals in this study. For each sample, the PhIP-Seq enrichment was compared to each of the 78 other samples. We found that individual autoreactomes were distinctive, with very little similarity to others (Pearson R-coefficient median = 0.021; Q1=0.018, Q3=0.023)(Fig. 1a: Right Panel).

### The autoreactome is longitudinally stable

While individual autoreactomes appear distinct, it remained an open question whether these profiles were stable over time within a given individual. To address this question, we performed PhIP-Seq on longitudinal serum samples from 7 distinct healthy individuals collected over a median of 63 months (35 samples total; Q1=55.5 months, Q3=76 months) (“Longitudinal” demographics in Extended Data Table 1). None of these individuals were being treated with immunomodulatory agents at any point during sample collection, however one had basal cell carcinoma and another had a history of Sjögren’s disease (clinical details in Extended Data Table 2). We compared the complete PhIP-Seq enrichment profile for each sample to all other samples. (Fig. 1b). These results clearly revealed that the intraindividual autoreactome profiles were highly correlated (Pearson R-coefficient median=0.883; Q1 =0.817, Q3=0.940). Conversely, the autoreactome of longitudinal samples within an individual was significantly more similar to each other than to the autoreactomes between different individuals (Mann-Whitney U p-value=4.91e^−95^). Additionally, the distribution of intraindividual correlations overlapped considerably with the distribution from technical replicates (104 of 146 (71.2%) longitudinal sample R-values fall within 2 standard deviations of the mean of technical replicate R-values), indicating that in most cases longitudinal autoreactomes are as similar to one another as technical replicates (Fig. 1c). These results suggest that the dominant humoral determinants of autoreactivity within an individual are not subject to large variation by this assay, at least within the median 5-year time scale of this analysis.

### The autoreactome is driven by rare autoreactivities and minimally altered by IVIG

We and others have used PhIP-Seq to investigate autoimmune disease determinants; however, many patients with immune disorders or deficiencies are treated with intravenous immunoglobulin (IVIG). IVIG is pooled from thousands of donors^35^ and therefore may contain relatively common autoantibodies, which has the potential to confound autoantibody assays^36^. To investigate the effect of IVIG on intraindividual PhIP-Seq performance, we examined a cohort of 189 samples from patients with myasthenia gravis, an autoantibody-mediated autoimmune disease. Among these 189 samples we identified 4 paired sets of samples in which the first collection was in a patient naïve to any immunomodulatory treatments, the second sample was within 6 weeks of IVIG treatment (2 within days, 1 within 3 weeks, 1 within 6 weeks), and no additional immunomodulatory treatments had been given except for steroids and in one case azathioprine (“IVIG” demographics in Extended Data Table 1; sample details in Extended Data Table 3).

PhIP-Seq was performed on these samples, and the mean correlation before and after IVIG was 0.815 (relative to 0.87 in longitudinal samples from individuals over time without any intervention)(Extended Data Fig. 1a,b). To further determine whether there were directional differences in the levels of PhIP-Seq detected autoantibodies following IVIG treatment, the sum of the top 10 differentially enriched autoantibodies (see “Methods) derived from each individual before and after IVIG administration was compared. No significant difference was observed (two-sided paired-samples Wilcoxon test p-value=0.625) (Extended Data Fig. 1c) before and after IVIG.

### Rituximab treatment has minimal effect on the autoreactome

Depletion of CD20 positive B cells with rituximab is a common treatment in autoimmunity and presumed autoantibody mediated diseases^37–39^. To determine the extent to which rituximab treatment alters the autoreactome, we examined our cohort of 189 samples from patients with myasthenia gravis to identify pairs of pre- and post-rituximab treatment samples. Like other autoantibody mediated diseases, treatment of myasthenia gravis can include multiple concurrent therapies which could potentially alter the autoantibody profile of an individual. To be conservative, we excluded all patients who had received any immunomodulation (including IVIG and plasma exchange) other than rituximab, steroids, or azathioprine, and for whom a pre-treatment sample was not available. Using these stringent criteria, 35 longitudinal samples were identified from 7 individuals and analyzed by PhIP-Seq (“Rituximab” demographics in Extended Data Table 1; sample clinical details in Extended Data Table 4).

The PhIP-Seq enrichment profile for each sample from a given individual who received rituximab was compared. Despite rituximab therapy, the autoreactome remained overall stable within each individual over time (Pearson R-coefficient mean of 0.887; Q1=0.782, Q3=0.940)(Fig. 2a). The overall distribution of correlation coefficients from individuals over time who received rituximab was similar to individuals who did not receive rituximab (108 of 136 (79.4%) within 2 standard deviations of the mean of R-values from longitudinal samples without interventions) and was not significantly different (Mann-Whitney U test p-value=0.66)(Fig. 2b). While rituximab is known to transiently reduce certain antibodies, these results suggest the overall profile of autoreactivity post rituximab treatment remains essentially unchanged.

**Figure 2.**
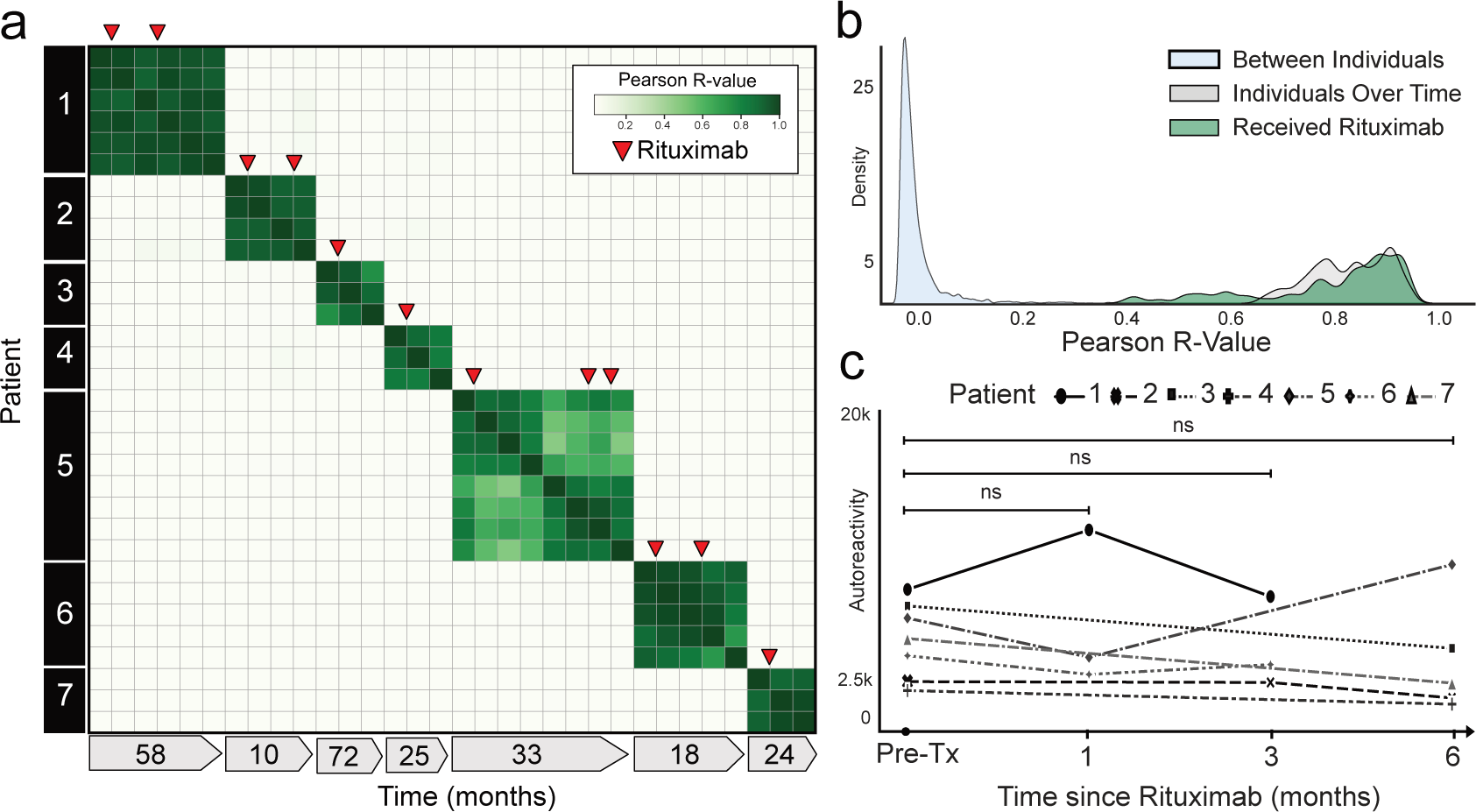
Rituximab treatment does not significantly alter the autoreactome. **(a)** Correlation matrix showing Pearson R-values of complete PhIP-Seq signal in 7 distinct individuals with myasthenia gravis each of whom were either rituximab naive or had not received rituximab for more than 6 months prior to first sample collection. Red arrows represent administration of rituximab. **(b)** Kernel density estimate plot showing distribution of Pearson R-value correlation coefficients among longitudinal samples from individuals receiving rituximab relative to longitudinal samples from individuals received no intervention. **(c)** Lineplots showing the autoreactivity (sum of top 10 PhIP-Seq Z-scores relative to the 79 healthy controls) for each patient over the first 6 months following initial rituximab dose. One-way paired samples Wilcoxon test p-value=0.625 at 1 month, 0.125 at 3 months, and 0.3125 at 6 months.

To determine whether there were decreases in subsets of PhIP-Seq enriched autoreactivities following rituximab treatment, as opposed to the complete profile, the sum of the top 10 differentially enriched protein targets (see “Methods”) in each individual were calculated at the time of initial sample collection, and then tracked longitudinally following the first dose of rituximab. To avoid overlapping timelines, datapoints following an additional round of rituximab therapy were removed from this analysis. There was no significant difference in the overall autoreactivity at 1, 3, or 6 months post-rituximab therapy (one-way paired samples Wilcoxon test p-value=0.625 at 1 month, 0.125 at 3 months, and 0.3125 at 6 months) (Fig. 2c). While PhIP-Seq enrichment does not report on absolute immunoglobin levels, the levels of the disease-causing autoantibody (either anti-AChR or anti-MuSK antibodies) in myasthenia gravis were measured independently by a clinical radioimmunoassay (RIA; either Athena Diagnostic or Mayo Clinic Laboratory) in 6 of the 7 patients at the same timepoints. Although autoantibody levels minimally decreased in 3 patients, and moderately decreased in the other 3 patients following rituximab treatment, they never fell below the established positive cutoff for the assay (Extended Data Fig. 2), suggesting that rituximab therapy was unable to quantitatively remove the pathogenic autoantibodies.

### CD19+ B cells are not required to maintain the autoreactome

To evaluate the impact of CD19 positive B cell depletion on the autoreactome, PhIP-Seq was performed on samples prior to, and approximately 6-months following, anti-CD19 CAR-T therapy in 14 individuals being treated for lymphoma (“CD19 CAR-T” demographics in Extended Data Table 1) who achieved and remained in remission, indicating successful depletion of the targeted cells. In 13 of the 14 patients, circulating CD19 cells were either persistently absent (defined as less than 10 CD19+ B cells per microliter) or became absent following treatment. In the remaining patient, only a sample prior to therapy was available, and CD19 B cells were already absent, indicating that they likely remained absent following additional targeted CD19 depleting therapy (Extended Data Fig. 3). None of the individuals had received an allogeneic hematopoietic cell transplant (HCT) in the preceding year (though two had received an autologous HCT), and 12 of the 14 patients were free of any additional B cell depleting therapies for the 6 months prior to initial sample collection. Intravenous immunoglobulin (IVIG) is often administered to patients receiving B cell depleting therapy, and 5 of the 14 patients received IVIG (”CD19” sample clinical details in Extended Data Table 5).

The complete PhIP-Seq enrichment profile within each individual before and after CD19 CAR-T therapy was once again compared. Despite depletion or persistently absent CD19 B cells in the setting of active CD19 CAR-T therapy, the autoreactome remained remarkably stable over time (Pearson R-coefficient median = 0.850; Q1=0.770, Q3=0.921) (Fig. 3a).

**Figure 3.**
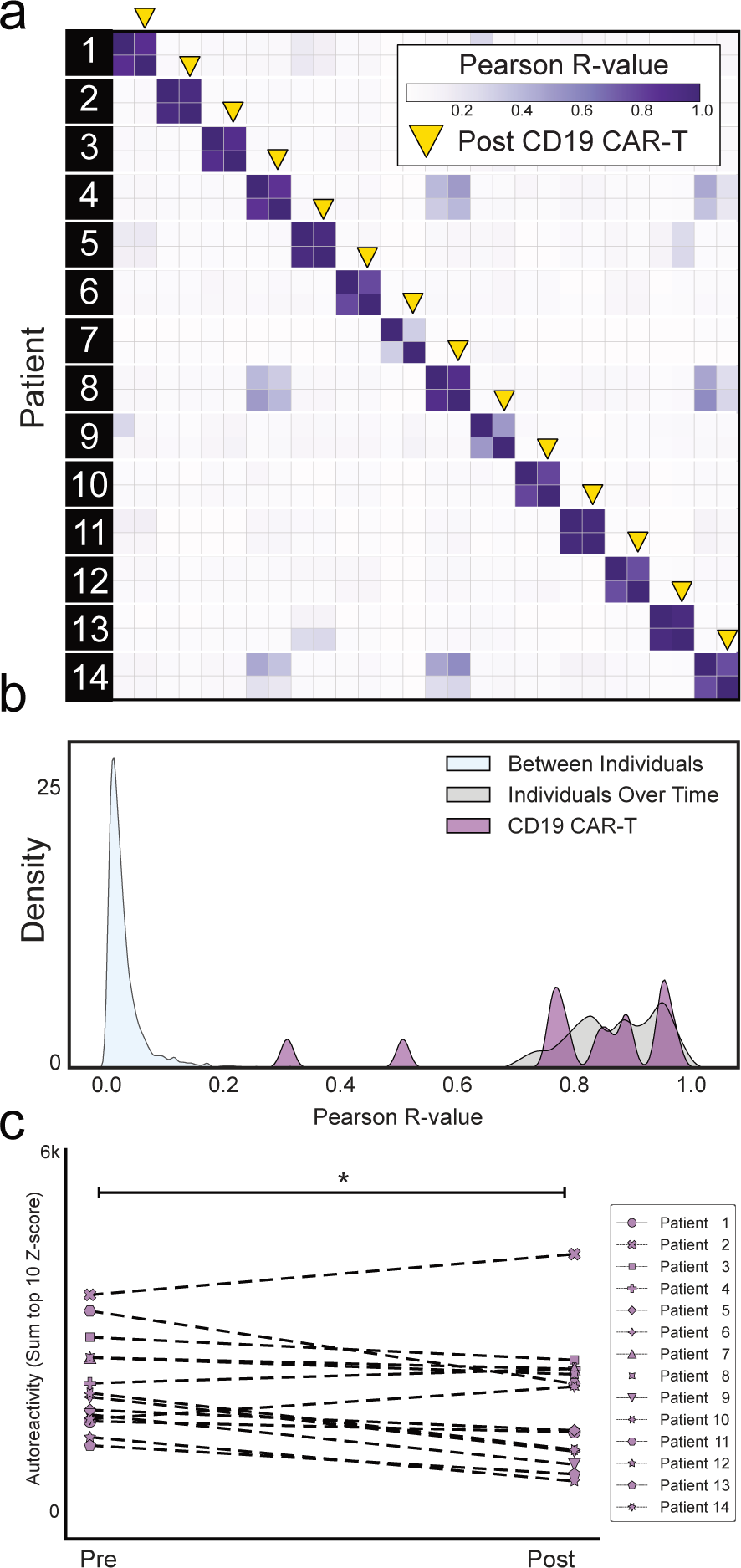
CD19 CAR-T therapy has minimal effect on the autoreactome 6 months after treatment. **(a)** Correlation matrix showing Pearson R-values of complete PhIP-Seq signal in 14 distinct individuals before and after anti-CD19 CAR-T therapy. Yellow arrows represent the 6-month post-treatment timepoint. **(b)** Kernel density estimate plot showing distribution of correlation coefficients within each individual before and after therapy relative to the distribution among untreated individuals over time and between different individuals. **(c)** Lineplots showing the autoreactivity (sum of top 10 PhIP-Seq Z-scores relative to the 79 healthy controls) for each patient before and after treatment. One-sided paired samples Wilcoxon test p-value=0.021.

The overall distribution of correlation coefficients in the CD19 CAR-T therapy group was not significantly different from intraindividual variation (Mann Whitney U p-value=0.284) The distribution of correlation values in 12 of the 14 (85.7%) patients fell within the distribution (2 standard deviations from the mean) of longitudinal samples from healthy individuals who received no interventions (Fig. 3b).

Using the same analysis approach as described for rituximab, we assessed differential PhIP-Seq enrichments using the sum of top 10 most enriched protein targets before and after CD19 CAR-T treatment (see “Methods”). Of the 14 individuals evaluated, 11 had a decrease in enrichment values while the remaining 3 had an increase, and levels overall were significantly decreased (one-sided paired samples Wilcoxon test p-value=0.021) (Fig. 3c). However, the size of the effect was minimal (median percent decrease in autoreactivity of 11.9%). These results suggest that similar to rituximab, sufficient immunoglobin producing cells remain after treatment such that the pattern of autoreactivity by PhIP-Seq remains largely unaltered.

To orthogonally validate the PhIP-Seq results, serum from 9 of these 14 patients were assayed using a previously described, multiplexed micro-bead assay consisting of 55 known protein autoantigens, each of which was covalently bound to microbeads with distinct barcodes^40^. The list of autoantigens included proteins targeted in connective tissue diseases such as SLE, scleroderma, and myositis, as well as secreted proteins such as cytokines, chemokines and growth factors. Additionally, antibody signal to 21 viral antigens was tested to determine whether antibodies targeting both self-proteins and viral proteins respond similarly in the absence of CD19+ B cells (see Extended Data Table 6 for a list of antigens). Because IVIG contains autoantibodies that confound measurements in bead-based assays (our unpublished observations), we excluded all patients who had received IVIG within 8 weeks of the initial blood draw (pre CAR-T), or had received interim IVIG between the pre CAR-T and post CAR-T (6 months after) blood draw. Nine of our 14 patients met these stringent criteria and were included in these experiments.

As expected, a minority of autoantigens were recognized by serum IgG autoantibodies, including 3 intracellular proteins (thyroperoxidase, TPO; bactericidal permeability inducing protein, BPI; and pyruvate dehydrogenase complex (PDC)) and 13 secreted proteins. Although levels of fourteen of these 16 autoantibodies had decreased signal overall (sum of normalized MFIs; see “Methods”) following anti-CD19 treatment, this decrease was only statistically significant in one case (Extended Data Fig. 4a). Among the 17 antiviral antibodies with meaningful signal, 12 were lower following CD19 therapy, but none were statistically significant (Extended Data Fig. 4b). These data, generated with an orthogonal platform using full-length proteins as targets confirms that the autoreactome, as well as IgG responses to viruses, remain largely stable over time following anti-CD19 CAR-T therapy.

### The autoreactome is profoundly altered following depletion of BCMA positive B cells

While CD19 is known to be expressed on a subset of antibody secreting plasma cells^29^, BCMA is a marker expressed on all plasma cells^31^, making it an attractive target for broad autoantibody depletion. To assess the impact of BCMA CAR-T on the autoreactome, we performed PhIP-Seq on serum samples from 9 individuals before, and approximately 6-months following, successful treatment with BCMA targeted CAR-T therapy (“BCMA” demographics in Extended Data Table 1). All 9 individuals had confirmed depletion of plasma cells in bone marrow following anti-BCMA CAR-T treatment (Extended Data Fig. 5). Each individual was being treated for multiple myeloma, none had received a HCT in the previous year, and 6 of the 9 had not received any additional B cell depleting therapy in the prior year. All post-treatment samples were collected at least 56 days from the last dose of IVIG, and 3 of the 9 patients never received interim IVIG (“BCMA” sample clinical details in Extended Data Table 5).

The complete PhIP-Seq enrichment profile obtained for each individual before and after anti-BCMA CAR-T therapy was compared. In contrast to CD19 and CD20 targeting therapies, the autoreactome was essentially devoid of any similarity following BCMA targeted therapy (Pearson R-value median = 0.006; Q1=0.002, Q3=0.130) for 8 of the 9 individuals (Fig. 4a). The autoreactome of one individual remained unaltered (R-value = 0.894). This individual was subsequently found to have relapsed around the time of sample acquisition, indicating potential failure of the CAR-T treatment. The overall distribution of correlation coefficients in the BCMA CAR-T therapy group was significantly different from alterations in healthy individuals over time without interventions (Mann Whitney U p-value=0.000012)(Fig. 4b). The samples from the individual with disease relapse were the only set whose autoreactome remained within 2 standard deviations from the mean of individuals over time without treatment. Remarkably, of the remaining 8 individuals, 7 of them had autoreactome correlation values following anti-BCMA CAR-T therapy that fell within 2 standard deviations of the mean of samples taken from entirely different individuals. The observed complete “reset” of the autoreactome in these patients suggest that successful treatment with anti-BCMA CAR-T cells sufficiently removes a lifetime of accumulated antibody producing plasma cells.

**Figure 4.**
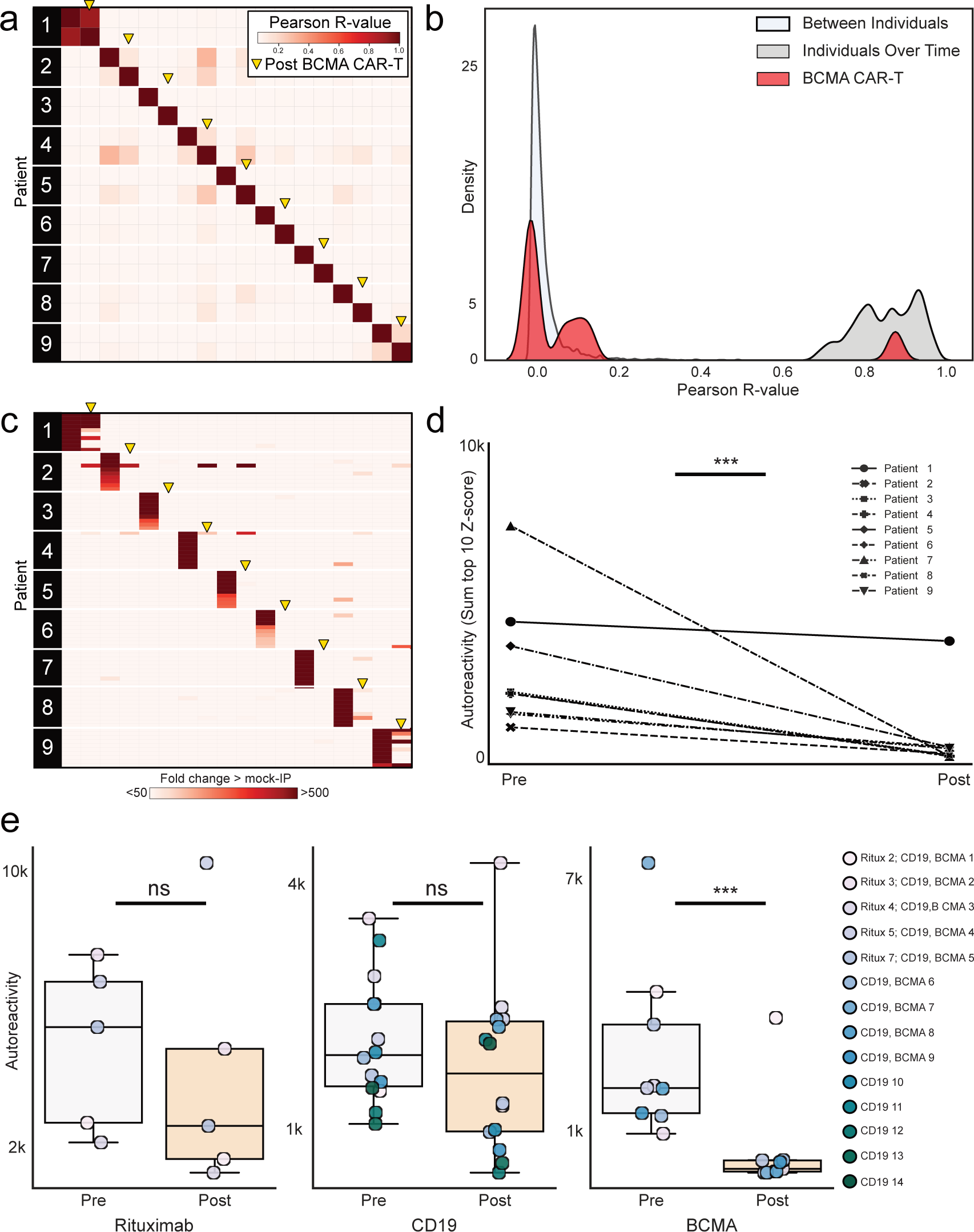
Anti-BCMA CAR-T therapy significantly alters the autoreactome. **(a)** Correlation matrix showing Pearson R-values of complete PhIP-Seq signal in 9 distinct individuals before and after anti-BCMA CAR-T therapy. Yellow arrows represent 6 month post-treatment time point. **(b)** Kernel density estimate plot showing distribution of correlation coefficients within each individual before and after BCMA-targeted therapy relative to the distribution among untreated individuals over time, and the distribution between different individuals. **(c)** Heatmap showing top 10 autoreactivities (sum of top 10 PhIP-Seq Z-scores relative to the 79 healthy controls) in each individual before and after treatment. **(d)** Lineplots showing the autoreactivity (sum of top 3 through 12 PhIP-Seq Z-scores relative to the 79 healthy controls therefore accounting for potential paraprotein confounding) for each patient before and after treatment. One-sided paired samples Wilcoxon test p-value=0.0019. **(e)** Boxplots showing the relative distributions of autoreactivity before and after treatment with rituximab (sum of top 10), anti-CD19 CAR-T (sum of top 10), and anti-BCMA CAR-T (sum of top 3 through 12 autoreactivities). Rituximab treatment cohort Mann-Whitney U p-value=0.42 with a median percent decrease of 52.3%; anti-CD19 CAR-T treatment cohort Mann-Whitney U p-value=0.206 with a median percent decrease of 11.9%; rituximab treatment cohort Mann-Whitney U p-value=0.003 with a median percent decrease of 95.6%.

While the overall autoreactive profile in sera following anti-BCMA CAR-T treatment is markedly altered, we also examined changes to the most differentially enriched protein targets from pre-treatment samples. Patients with multiple myeloma have a monoclonal expansion of a single plasma cell which secretes paraprotein antibody. Because paraprotein is potentially autoreactive, and because it is known to dramatically decrease following BCMA CAR-T treatment in multiple myeloma, we removed the top two enriched protein targets from each individual prior to analysis to minimize the chance that our results were being confounded by changes in paraprotein level. The autoreactivity levels for the top 3-12 proteins decreased in every patient follow anti-BCMA CAR-T therapy, and the overall change was statistically significant (one-sided paired samples Wilcoxon test p-value=0.0019) (Fig. 4c and 4d). Unlike anti-CD19 CAR-T treatment, in which the size of the decrease was minimal, there was a 95.6 percent decrease in PhIP-Seq enrichment following anti-BCMA CAR-T treatment (Fig. 4e). To ensure these findings were similar if the top 10 protein targets were used (and potential confounding by paraprotein is not accounted for), the same analysis was performed without the removal of the top 2 autoreactivities, with similar findings (Extended Data Fig. 6).

To orthogonally validate the PhIP-Seq results and further explore the effects of plasma cell depletion on antiviral antibodies, we used the same multiplexed bead-based arrays described earlier to characterize IgG binding to 55 autoantigens and 21 viral proteins. However, because IVIG is routinely given following BCMA CAR-T therapy, only 3 of the original 9 patients were IVIG free, so samples from 4 additional patients meeting the stringent criteria outlined previously were used (demographics and sample details in Extended Data Tables 7 and 8).

Of the same 16 measurable autoantigens analyzed in the anti-CD19 CAR-T treatment cohort, 13 had overall decreased IgG binding (sum of normalized MFIs; see “Methods”) following anti-BCMA CAR-T treatment. In contrast to the CD19 CAR-T cohort in which there was only one autoantibody with a statistically significant decrease, levels of 9 of the 16 measured autoantibodies significantly decreased following anti-BCMA CAR-T therapy (Extended Data Figure 7a). Also, unlike the CD19 CAR-T cohort in which levels of none of the anti-viral antibodies significantly decreased, all 17 of the measured antiviral antibodies decreased following BCMA therapy, and 8 of these decreases were statistically significant (Extended Data Figure 7b).

## Discussion

Adaptive immune responses targeting self rather than foreign proteins are a hallmark of autoimmune disease. Previous work has noted the existence of circulating self-reactive antibodies in healthy individuals. This work has focused on “natural autoantibodies”, which are thought to arise without antigen stimulation and to consist primarily of unmutated, polyreactive, often IgM-class antibodies^41,42^. Despite considerable interest, the role of these antibodies in health and disease remains unclear, though there is evidence of variable and context-dependent roles; the autoantibodies can contribute to, or protect against, inflammatory and autoimmune disease^43–53^. The natural autoantibody repertoire is thought to be relatively shared amongst individuals, and to develop during the first few years of life^42,54–56^. Though natural autoantibodies may target autoantigens present in autoimmune disease, substantial evidence exists that the specific epitope differs from the disease-associated autoantibody, which may in part explain why the healthy individuals studied here are clinically immune tolerant ^57,58^.

Here, through the targeted application of a custom proteome-wide autoantibody detection technology, we find that the repertoire of auto-reactive immunoglobins present in each healthy individual is highly distinctive. This “autoreactome” is described by the complex set of thousands of peptides reproducibly enriched by immunoprecipitation, and apparently unique to each individual. We observe that the autoreactome profile of individuals is remarkably stable, even over years of time, and thus may be considered a “serological fingerprint.” In contrast to natural autoantibodies, the autoreactome is characterized by uniqueness in each individual, rather than a common, shared set of autoreactivities. Interestingly, the stability and uniqueness of the autoreactome suggests that our approach may be utilized to de-convolute the origin of an unknown patient serum sample in repositories where multiple samples have been collected at different timepoints from the same patient.

During a humoral immune response, plasma cells of varied lifespans are generated, with short-lived plasmablasts providing rapid antibody production in the acute phase and long-lived plasma cells sustaining durable antibody titers. While foreign antigen-specific long-lived plasma cells are known to persist for decades in humans^59–61^, the cellular phenotype of pathogenic plasma cells remains controversial. Since individual autoantibodies decline following therapeutic B cell ablation, despite a lack of CD19 and CD20 expression on plasma cells^27,62,63^, the prevailing model holds that the continuous generation of short-lived plasma cells from memory B cell precursors underlies the autoantibody repertoire^64^. In contrast with this view, we show that not only is the broader intraindividual autoreactome stable over time, but that this autoantibody repertoire is resistant to therapeutic B cell ablation with rituximab (anti-CD20) and anti-CD19 CAR-T cells. Given the expanded options for targeting B cells and plasma cells in systemic autoimmunity, this enhanced understanding of autoreactome biology has important clinical implications. Indeed, by using the autoreactome as a proxy for the autoantibody repertoire within an individual, we show here that depletion of plasma cells via anti-BCMA CAR-T therapy drastically alters the autoantibody repertoires. In most cases, the autoreactome of a sample after treatment with BCMA CAR-T cells appears as similar to an entirely different person as to themselves prior to treatment.

It has been shown that rituximab reduces the level of certain autoantibodies^65^, and has clinical efficacy in many autoimmune disease settings, which may relate more to the antigen presentation function of CD20-positive B cells as proposed by others^66^. Additionally, CD19 CAR-T therapy has recently been shown to be effective in the treatment of refractory SLE. Because both anti-CD19 CAR-T and anti-BCMA CAR-T are primarily used in the treatment of hematologic malignancy, our cohorts received chemotherapy prior to CAR-T therapy, and therefore had low CD19 and plasma cells numbers even prior to the initiation of therapy. Future studies will be required to further understand the relationship of the pre- and post-CAR-T treatment autoreactomes, and its relationship to clinical efficacy, in the specific context of autoimmune diseases. Our broad based approach to screen autoantibodies at a proteome wide level allows a more comprehensive and unbiased assessment of the effect of therapies on the specificity of the autoreactome. We posit that this approach could greatly complement emerging data on the use of CAR-T cell therapies that are being utilized against severe autoimmune diseases.

The prevalence and burden of autoimmune disease continues to rise^67,68^, yet the precise mechanisms for the development of autoreactivity remain unclear. There is mounting evidence that combinations of genetic and environmental risk factors, including viral infections, predispose individuals to disease. In addition to the emerging therapies already discussed, other new therapies exist which can halt or prevent the progression of certain autoimmune diseases, including type 1 diabetes (T1DM)^69^. By defining the autoreactome and describing the expected longitudinal changes over time, we introduce a new tool which can be applied for additional basic and translational studies in autoimmunity. Future work will be needed to explore the autoreactome to determine at what age it develops, whether there are shared features of autoreactive epitopes, what drives the development of the autoreactivities, and what biological role these autoreactive antibodies may have in health and disease. Beyond the already discussed application for evaluating the relative effectiveness of various emerging therapies, there are additional potential future applications of this work by tracking longitudinal autoreactome changes in individuals prior to, and following, the development of specific autoimmune diseases. This may allow for the identification of autoreactive signatures which precede the development of symptoms, and thereby produce novel biomarkers to identify patients in whom autoimmune disease could be prevented.

## Data Availability

All PhIP-Seq data will be made publicly available upon publication of this manuscript via a Dryad digital repository; DOI: 10.5061/dryad.w3r2280z6

https://dx.doi.org/10.5061/dryad.w3r2280z6

## Methods

### Patients

The “Healthy Control” cohort consists of plasma from 79 self-reported healthy, pre-COVID-19 individuals which were obtained as deidentified samples from the New York Blood Center. These samples were part of retention tubes collected at the time of blood donations from volunteer donors who provided informed consent for their samples to be used for research.

For the bead-based protein arrays, serum and/or plasma was used for internal controls and validations, as has been previously described, as deidentified samples^70^. Negative controls included a pediatric sample from an infant between 6 and 12 months of age which was a gift from the Pulendran lab, as well as samples from 3 healthy subjects with known CMV-seronegative status confirmed by ELISA from the Stanford Blood Center. Positive controls include plasma samples derived from participants with autoimmune or other types of diseases with known reactivity patterns (e.g. thyroid peroxidase [TPO], pyruvate dehydrogenase complex [PDC-E2], fibroblast growth factor 7 [FGF7], proteinase 3, IL-11, CXCL-13) which were purchased from ImmunoVision or were obtained from Stanford Autoimmune Diseases Biobank and Oklahoma Medical Research Foundation (Oklahoma City, Oklahoma, USA; a gift of Judith James, Oklahoma Medical Research Foundation). A total of 12 positive control patient samples were used. All 16 of these patients (4 negative controls and 12 positive controls) provided informed consent for their samples to be used for research.

Longitudinal plasma samples were obtained from a set of 7 community dwelling, cognitively unimpaired, pre-COVID-19, healthy older adults recruited through the Brain Aging Network for Cognitive Health (BrANCH) at the University of California, San Francisco. All participants are screened at baseline and enrollment criteria excluded individuals with severe psychiatric illness, neurologic disorders (e.g., epilepsy, multiple sclerosis), and medical conditions that could impact cognition (e.g., recent substance use disorders, active chemotherapy). Following a comprehensive neurobehavioral evaluation, participants were classified cognitively normal per consensus conference with board-certified neurologists and neuropsychologists. Inclusion in the current study was contingent on no known autoimmune conditions at baseline or follow up visits, either active or inactive at the time of plasma collection. Study procedures were approved by the UCSF Committee on Human Research and all participants provided written informed consent (IRB #10-02076).

For the myasthenia gravis patient cohort (used to evaluate the effects of IVIG and rituximab), deidentified serum samples from myasthenia gravis patients, with laboratory confirmed AChR or MuSK autoantibody serostatus, were retrieved from a biorepository established at the Yale School of Medicine, Myasthenia Gravis Clinic (EXPLORE-MG Registry, NCT03792659) under the approval of Yale University’s Institutional Review Board.

The CAR-T cell cohorts consisted of adults aged ≥18 years with relapsed or refractory CD19+ or BCMA+ hematologic malignancies who received lymphodepleting chemotherapy with cyclophosphamide and fludarabine followed by anti-CD19 or anti-BCMA-CAR-T cells at Fred Hutchinson Cancer Center (Fred Hutch). Serum and PBMCs were prospectively collected once prior to lymphodepleting chemotherapy and at approximately 6-12 months after CAR-T cell infusion among individuals who achieved durable remissions and received no subsequent anti-tumor therapies. This work was approved by the Fred Hutch institutional review board (Protocol 10080), and all participants provided informed consent in accordance with the Declaration of Helsinki.

### PhIP-Seq

All PhIP-Seq was performed similar to our previously published multichannel protocol, with minor adjustments as outlined in our new protocol: https://www.protocols.io/view/derisi-lab-phage-immunoprecipitation-sequencing-ph-czw7×7hn?step=14.1

As previously described^11^, our human peptidome library consists of a custom-designed phage library of 731,724 unique T7 bacteriophage each presenting a different 49 amino-acid peptide on its surface. Collectively these peptides tile the entire human proteome including all known isoforms (as of 2016) with 25 amino-acid overlaps. 1 milliliter of phage library was incubated with 1 microliter of human serum overnight at 4C and immunoprecipitated with 25 microliters of 1:1 mixed protein A and protein G magnetic beads (Thermo Fisher, Waltham, MA, #10008D and #10009D). These beads were than washed, and the remaining phage-antibody complexes were eluted in 1 milliliter of E.Coli (BLT5403, EMD Millipore, Burlington, MA) at 0.5-0.7 OD and amplified by growing in 37C incubator. This new phage library was then re-incubated with the same individual’s serum and the previously described protocol repeated. DNA was then extracted from the final phage-library, barcoded, and PCR-amplified and Illumina adaptors added. Next-Generation Sequencing was then performed using an Illumina sequencer (Illumina, San Diego, CA) to a read depth of approximately 1 million per sample.

### Bead-based protein array

A 76-plex protein array comprising a 55-plex autoantigen protein subarray and a 21-plex viral protein subarray was constructed as previously described, with modifications^40^. In brief, antigens (Extended Data Table 6) were conjugated to uniquely barcoded carboxylated magnetic beads (MagPlex-C, Luminex Corp). The beads were stored in aliquots at −80°C after conjugation and thawed on the day of the experiment. 45 µl of diluted serum or plasma sample (1:100 in PBS-1%BSA) was transferred into the 384-well plate (Greiner BioOne) containing 5 µl of bead array per well. Samples were incubated for 60 min on a shaker at room temperature, and then at 4C overnight. The following day beads were washed with 4 × 60 µl PBS-Tween on a plate washer (EL406, Biotek) and then incubated with 50 µl of 1:1000 diluted R-phycoerythrin (R-PE) conjugated Fc-γ-specific goat anti-human IgG F(ab’)2 fragment (Jackson ImmunoResearch 109-116-170) for 30 min. The plate was washed with 4 × 60 µl PBS-Tween and re-suspended in 50 µl PBS-Tween prior to analysis using a FlexMap3D TM instrument (Luminex Corp.) and measuring MFI with at least 50 beads per barcode for each sample. Most of the individual antigens that had been conjugated to beads were qualified prior to mixing using antibodies-directed against epitope tags, monoclonal antibodies specific for the antigen, or prototype human plasma samples derived from participants with autoimmune diseases with known reactivity patterns (e.g., Scl-70+ systemic sclerosis; SSA+, which is associated with lupus and Sjögren’s syndrome; ribonucleoprotein (RNP)+ sera, which is associated with lupus and mixed connective tissue disease; and APS1 sera which are reactive with multiple cytokines; data not shown). Binding events were displayed as Median Fluorescence Intensity (MFI). MFI values were normalized by subtracting bare-bead MFI values. Replicate MFI values were averaged.

### Enumeration of B cells and plasma cells

B cells were quantified using a research flow cytometry panel (Representative gates shown in Extended Data Fig. 8). Briefly, peripheral blood mononuclear cells (PBMCs) were incubated on ice for 30 minutes with fluorescently labeled antibodies (in fluorescence-activated single cell sorting [FACS] buffer containing Dulbecco’s phosphate-buffered saline [DPBS] and 1% newborn calf serum [Life Technologies]) before its analysis on a FACSymphony A5 (BD Bioscience) flow cytometer. The following antibodies were included for cell labeling: fixable viability stain 700 (BD cat#564997), anti-CD19 BV421 (HIB19, BD), anti-CD45 BV510 (HI30, BD), anti-CD3 BV605 (UCHT1, BioLegend), anti-CD16 BV711 (3G8, BD), anti-CD14 BV711 (M0P-9, BD). FlowJoTM Software version 10.7.1 was used for analyses with flow cytometry proportions multiplied by absolute lymphocyte counts to calculate total CD19^+^ B cell numbers. Plasma cells in the bone marrow were quantified based on detection of CD138^+^ plasma cells in clinically obtained bone marrow core biopsies by immunohistochemistry in the University of Washington Hematopathology Laboratory.

### Analysis of PhIP-Seq

As previously described^11^, next generation sequencing reads from fastq files were aligned at the level of amino acids using RAPSearch2. All results represent the average of technical replicates, except for the 79 individual healthy controls in whom only 24 of the individuals (48 samples) were performed in technical replicate. All human peptidome analysis was performed at the gene-level, in which all reads for all peptides mapping to the same gene were summed, and 0.5 reads were added to each gene to allow inclusion of genes with zero reads in mathematical analyses. Within each individual sample, reads were normalized by converting to the percentage of total reads. To normalize each sample against background non-specific binding, a fold-change (FC) over mock-IP was calculated by dividing the sample read percentage for each gene by the mean read-percentage of the same gene for the AG bead only controls. This FC signal was then used for side-by-side comparison between samples and cohorts. FC values were also used to calculate z-scores for specific cohorts relative to defined controls.

### Analysis of multiplexed bead arrays

Antibody reactivity to each target for each sample was measured in mean fluorescent intensity (MFI). Data is only shown for targets which have an MFI of at least 1000 MFI units at any time point for any patient. To adjust for background binding to beads only, each sample was additionally run on unconjugated beads, and bare bead MFI was subtracted to create an adjusted MFI for each target antigen within the same corresponding sample. To account for nonspecific binding from human serum, a sample from an infant control known to lack antibodies to any of the viral or human targets was run in duplicate. The mean of the adjusted MFI’s for the control duplicates was calculated for each target, and further subtracted from each corresponding target from each sample. A normalization function was then performed such that all sample values for each target antigen ranged from zero to one, referred to as “Normalized MFI”.

### Statistical methods

All statistical analysis was performed in Python using the Scipy Stats package. To calculate the overall similarity of PhIP-Seq detected autoreactive profiles between different samples, a Pearson correlation coefficient was calculated using all FC over mock-IP values (unless specifically stated otherwise) for each autoantigen in each sample relative to each autoantigen in each other sample. For comparisons of distributions of PhIP-Seq signal (either autoreactivity scores or Pearson R-values) between two groups, a non-parametric Mann-Whitney U test was utilized. To determine whether specific interventions (IVIG, rituximab, anti-CD19 CAR-T, or anti-BCMA CAR-T) directionally decreased PhIP-Seq detected autoreactivities, first an “autoreactivity” score was calculated by summing the top 10 autoantigen z-scores (relative to the 79 healthy controls) in each pre-treatment sample. The sum of the z-scores from these same 10 autoantigens were again calculated following the intervention for each post-treatment sample from the same individual. These autoreactivity scores from before and after a treatment were paired within each individual, and used as input for a one-sided paired-samples Wilcoxon test. To determine whether anti-CD19 CAR-T treatment, or anti-BCMA CAR-T treatment altered Luminex detected antibody levels from the bead-based protein arrays, the pre-treatment and post-treatment normalized MFI values were compared using a two-sided paired-samples Wilcoxon test for each target antigen. P-values: ns > 0.05, * < 0.05, ** < 0.01, *** < 0.005, **** < 0.001.

## Acknowledgements

The authors acknowledge the New York Blood Center for contributing pre-COVID-19 healthy donor blood samples. We acknowledge Brian O’Donovan for coining the term “autoreactome”. The authors would like to thank Quinton R. Markett for assistance with organizing protein array reagents and design.

## Author Contributions

Conceptualization: A.B., H.L., M.R.W., J.A.H., S.W.J., M.S.A., J.L.D. Methodology: A.B., J.A., A.F.K., H.K., E.R., C.Y.W., A.S., P.J.U., A.R., M.K., M.S.A., J.L.D. Performed or contributed to experiments: A.B., D.J.L.Y., A.R., M.K., J.B., C.M.B., B.O. Formal analysis: A.B., J.L.D. Patient sample and clinical data acquisition: K.Z., K.D.D., V.E.D., E.Q.G., B.C., D.J.G, J.G., C.J.T., J.A.H., G.M., R.J.W, K.C.O. Clinical data curation: K.Z., K.D.D., K.B.C., J.H.K., V.E.D., E.Q.G., B.C., D.J.G, J.G., C.J.T., J.A.H., G.M, K.C.O. Writing (original draft): A.B., J.L.D. Writing (review and editing): A.B., K.C.O., P.J.U., J.A.H, S.W.J., M.S.A., J.L.D. Supervision: P.J.U., J.A.H, S.W.J., M.S.A., J.L.D.

## Competing Interest Declaration

### Funding

This work was supported by the Pediatric Scientist Development Program and the Eunice Kennedy Shriver National Institute of Child Health and Human Development grant K12-HD000850 (A.B.), and by the Chan Zuckerberg Biohub SF (J.L.D. and M.S.A.), and by National Institutes of Health grants R01AR073938 and R01AR075813 (S.W.J) and by and the National Cancer Institute (U01CA247548 to J.A.H.) and by the National Institute of Allergy and Infectious Diseases of the NIH under award numbers R01-AI114780(K.C.O), R21-AI142198(K.C.O), R21 AI164590 (K.C.O) and through an awards provided through award number U54-NS115054 (K.C.O) and NIH U54 NS115054 (G.M and R.J.N; G.M is also an MGNet Scholar Awardee) of the Rare Diseases Clinical Research Network Consortium (RDCRN) of the NIH and MGNet. All RDCRN consortia are supported by the network’s Data Management and Coordinating Center (DMCC) (U2CTR002818). This study was supported by NIH-NIA grants R01AG032289 (J.H.K) and R01AG072475(K.B.C), UCSF ADRC P30AG062422 (G.D.R), This work was also supported by a grant from the Larry L. Hillblom Foundation (2018-A-006-NET; J.H.K). This work was also supported by the Henry Gustav Floren Trust, the Stanford Department of Medicine Team Science Program, and funding from the Stanford Medicine Office of the Dean (P.J.U.).

### Potential Conflicts

J.D.R. reports being a founder and paid consultant for Delve Bio, Inc., and a paid consultant for the Public Health Company and Allen & Co. M.A.S. receives unrelated research funding from the NIH, the CDC, Cepheid and Merck and unrelated Honoria from UpToDate, Inc. M.R.W. reports being a founder and paid consultant for Delve Bio, Inc. and receives unrelated research grant funding from Roche/Genentech and Novartis, and received speaking honoraria from Genentech, Takeda, WebMD, and Novartis. C.J.T. reports being on the Scientific Advisory Boards for Caribou Biosciences, T-CURX, Myeloid Therapeutics, ArsenalBio, Cargo Therapeutics, Celgene/BMS Cell Therapy; is a member a DSMB member of Kyverna; is on Ad hoc advisory boards/consulting (last 12 months) for Nektar Therapeutics, Legend Biotech, Prescient Therapeutics, Century Therapeutics, IGM Biosciences, Abbvie; has Stock options in Eureka Therapeutics, Caribou Biosciences, Myeloid Therapeutics, ArsenalBio, Cargo Therapeutics; and reports the right to receive payment from Fred Hutch as an inventor on patents related to CAR T-cell therapy. J.A.H. reports research funding from Merck and Takeda and consulting fees from Takeda, Gilead, SentiBio, and Century Therapeutics. J.G. reports research funding from Sobi, Juno Therapeutics (a BMS company), Celgene (a BMS company), Angiocrine Bioscience; is an Ad hoc consultant for Sobi, Legend Biotech, Janssen, Kite Pharma, MorphoSys. D.J.G. has received research funding, has served as an advisor and has received royalties from Juno Therapeutics, a Bristol-Myers Squibb company; has served as an advisor and received research funding from Seattle Genetics; has served as an advisor for GlaxoSmithKline, Celgene, Janssen Biotech, Ensoma and Legend Biotech; and has received research funding from SpringWorks Therapeutics, Sanofi, and Cellectar Biosciences. K.C.O. received unrelated research funding from, and is an equity shareholder of Cabaletta Bio, and receives unrelated research funding from Seismic, argenx, and Viela Bio/Horizon (now Amgen). R.J.N. reports unrelated research support from the National Institutes of Health, Genentech, Inc., Alexion Pharmaceuticals, Inc., argenx, Annexon Biosciences, Inc., Ra Pharmaceuticals, Inc. (now UCB S.A.), the Myasthenia Gravis Foundation of America, Inc., Momenta Pharmaceuticals, Inc. (now Janssen), Immunovant, Inc., Grifols, S.A., and Viela Bio, Inc. (now Horizon Therapeutics plc). R.J.N. has also served as a consultant/advisor unrelated to research in this manuscript for Alexion Pharmaceuticals, Inc., argenx, Cabaletta Bio, Inc., Cour Pharmaceuticals, CSL Behring, Grifols, S.A., Ra Pharmaceuticals, Inc. (now UCB S.A.), Immunovant, Inc., Momenta Pharmaceuticals, Inc. (now Janssen), and Viela Bio, Inc. (now Horizon Therapeutics plc). J.L.D., C.M.B. and M.R.W. receive licensing fees from CDI Labs.

## Extended Data Figures and Tables

**Extended Data Figure 1.**
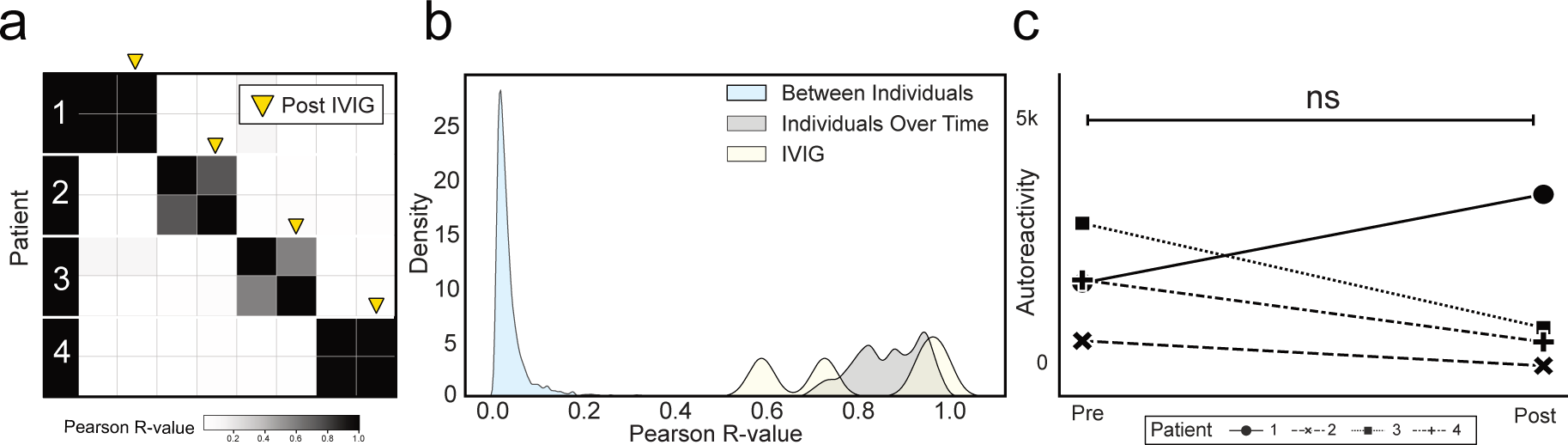
Administration of IVIG minimally alters the autoreactome. **(a)** Correlation matrices showing Pearson correlation coefficients of complete PhIP-Seq signal before and after IVIG administration. **(b)** Kernel density estimate plot showing distribution of Pearson R correlation coefficients from before and after IVIG administration relative to longitudinal samples from individuals over time, and between different individuals. **(c)** Lineplots showing the autoreactivity (sum of top 10 PhIP-Seq Z-scores relative to the 79 healthy controls) for each patient before and after IVIG administration. Paired-samples Wilcoxon test p-value 0.625.

**Extended Data Figure 2.**
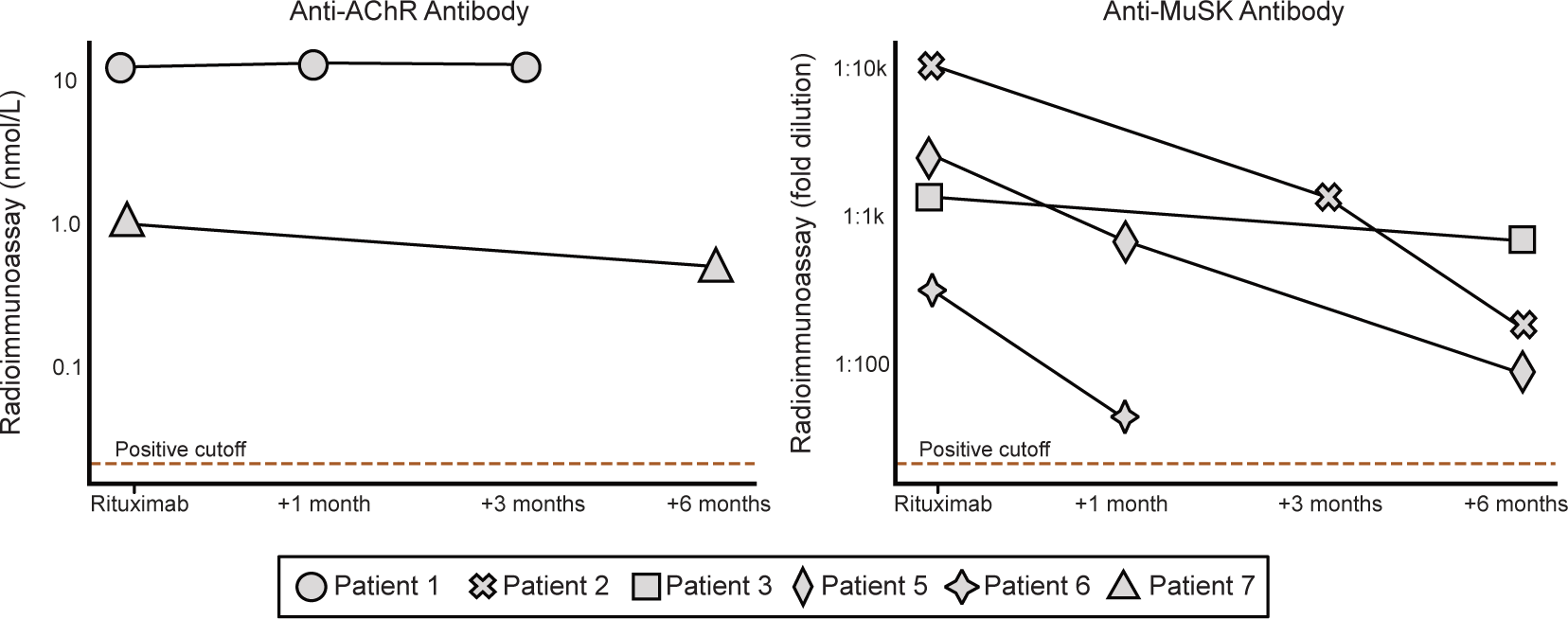
Changes in disease-causing autoantibody levels following rituximab Left. Anti-AChR autoantibody levels measured by radioimmunoassay (RIA; Mayo Clinic Laboratory, unit=nmol/L) in 2 patients at 1 month, 3 months, and 6 months post-rituximab. RIA positive cutoff 0.02 nmol/L. **Right**: Anti-MuSK autoantibody titers measured by RIA (Athenia Diagnostics, unit=fold dilution) in a different 4 patients at 1 month, 3 months, and 6 months post-rituximab. Positive cutoff for is a fold dilution of greater than 1:20.

**Extended Data Figure 3:**
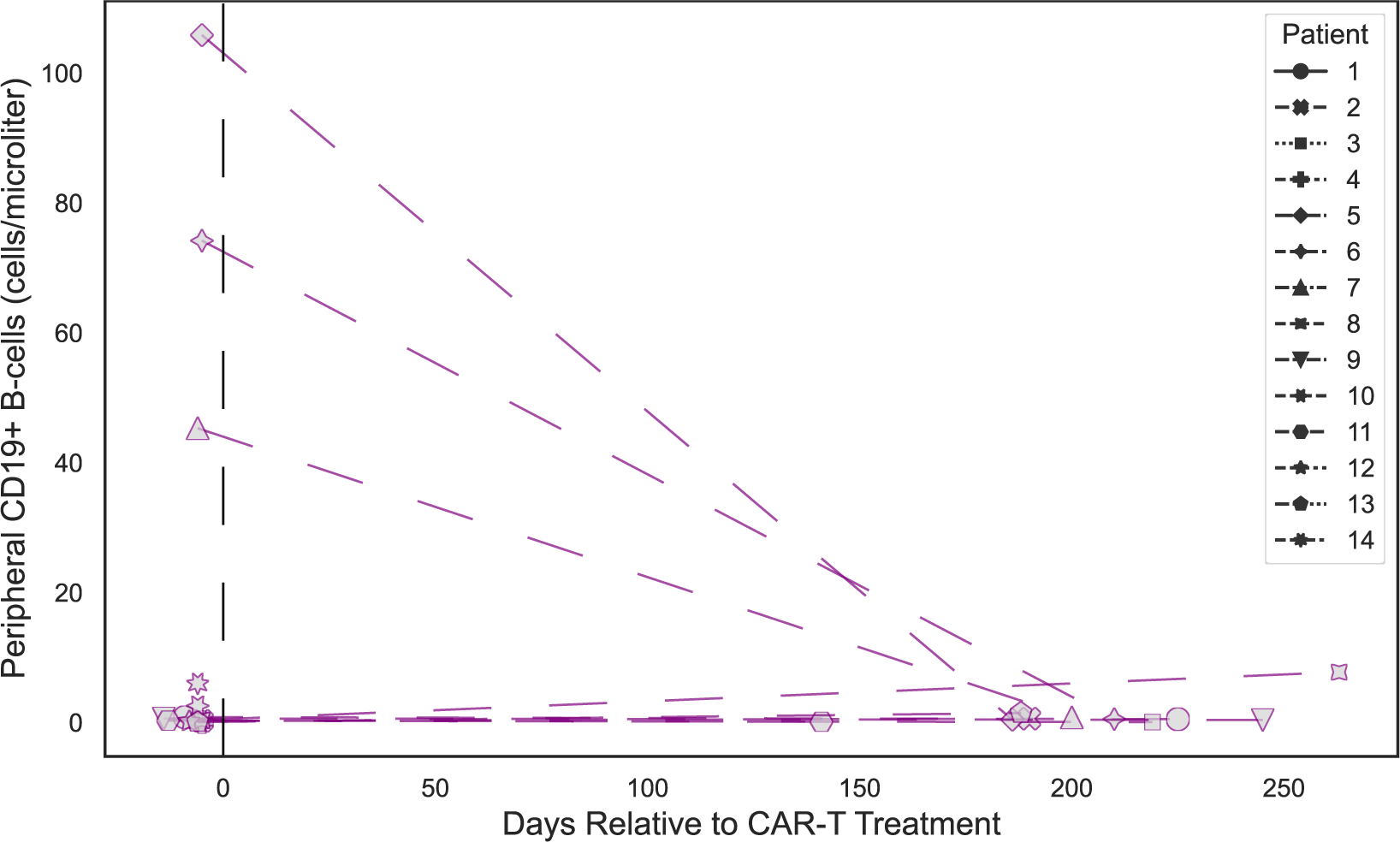
CD19 cell counts following anti-CD19 CAR-T treatment. Lineplots showing the levels of CD19^+^ B cells in peripheral blood measured by flow cytometry at various timepoints relative to anti-CD19 CAR-T treatment, in each patient.

**Extended Data Figure 4:**
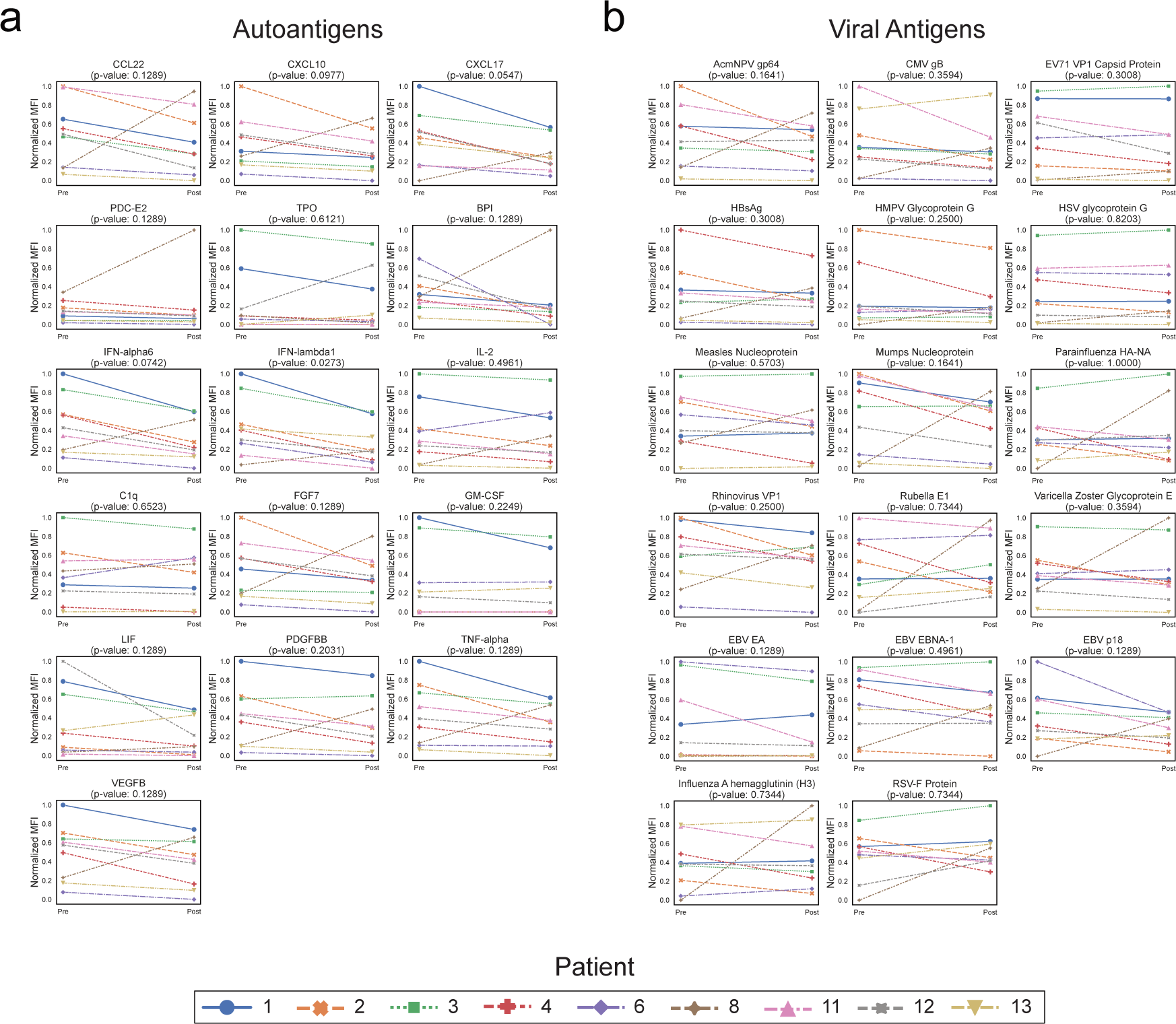
Changes in multiplex bead-based protein array detected antibody levels following CD19 therapy. **(a)** Multiplex bead-based protein array detected normalized MFI values (see “Methods”) for each of the 16 autoantigens with meaningful signal (see “Methods”) in each of the patients before and approximately 6 months after treatment with anti-CD19 CAR-T cells. P-values shown are from a paired-samples Wilcoxon test. **(b)** Multiplex bead-based protein array detected normalized MFI values (see “Methods”) for each of the 17 viral antigens with meaningful signal (see “Methods”) in each of the patients before and after treatment with anti-CD19 CAR-T cells. P-values shown are from a paired-samples Wilcoxon test.

**Extended Data Figure 5:**
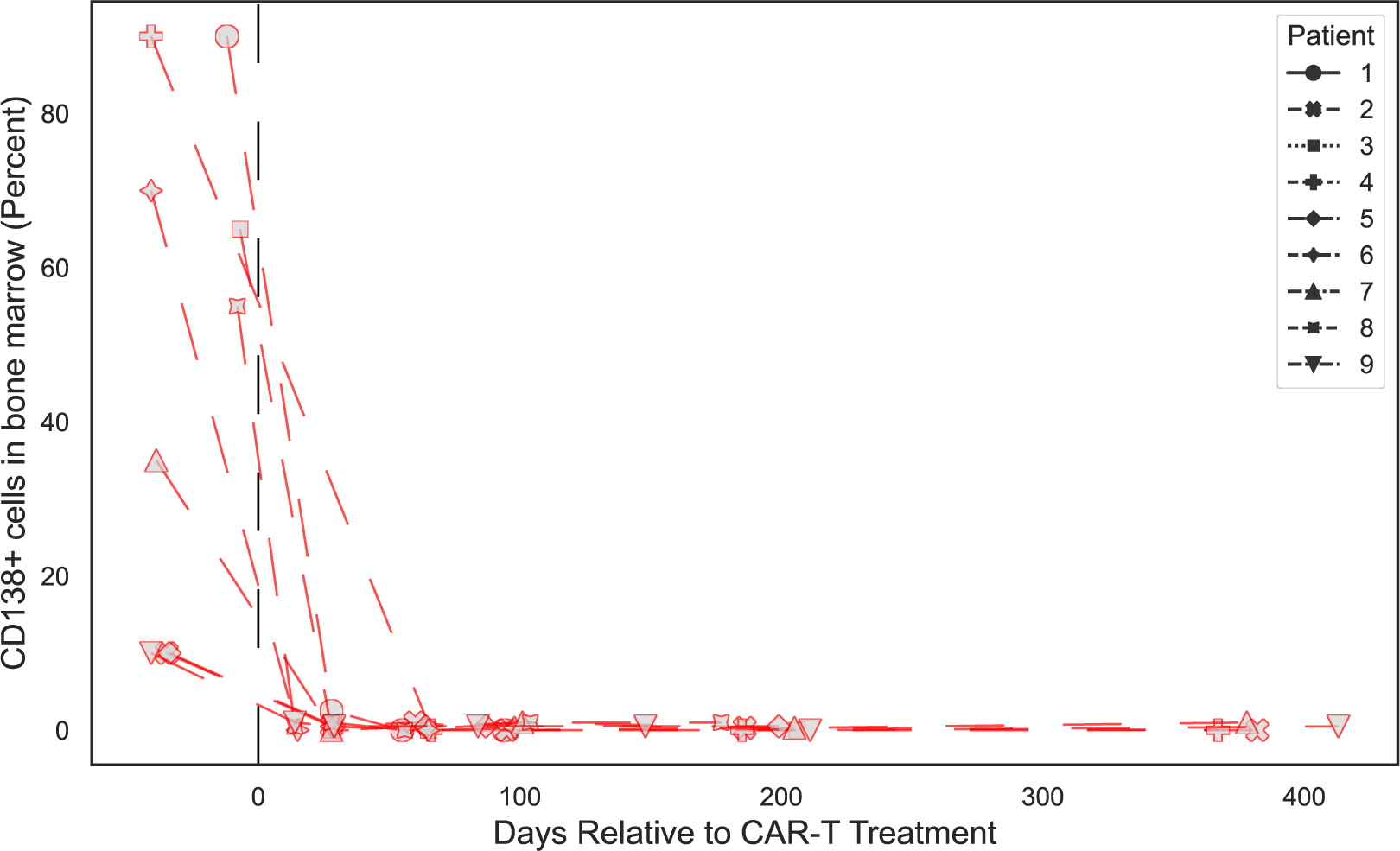
Changes in bone marrow plasma cell percentage following anti-BCMA CAR-T cell therapy. Lineplots showing the levels of plasma cells in the bone marrow measured by flow cytometry at various timepoints relative to anti-BCMA CAR-T treatment, in each patient.

**Extended Data Figure 6:**
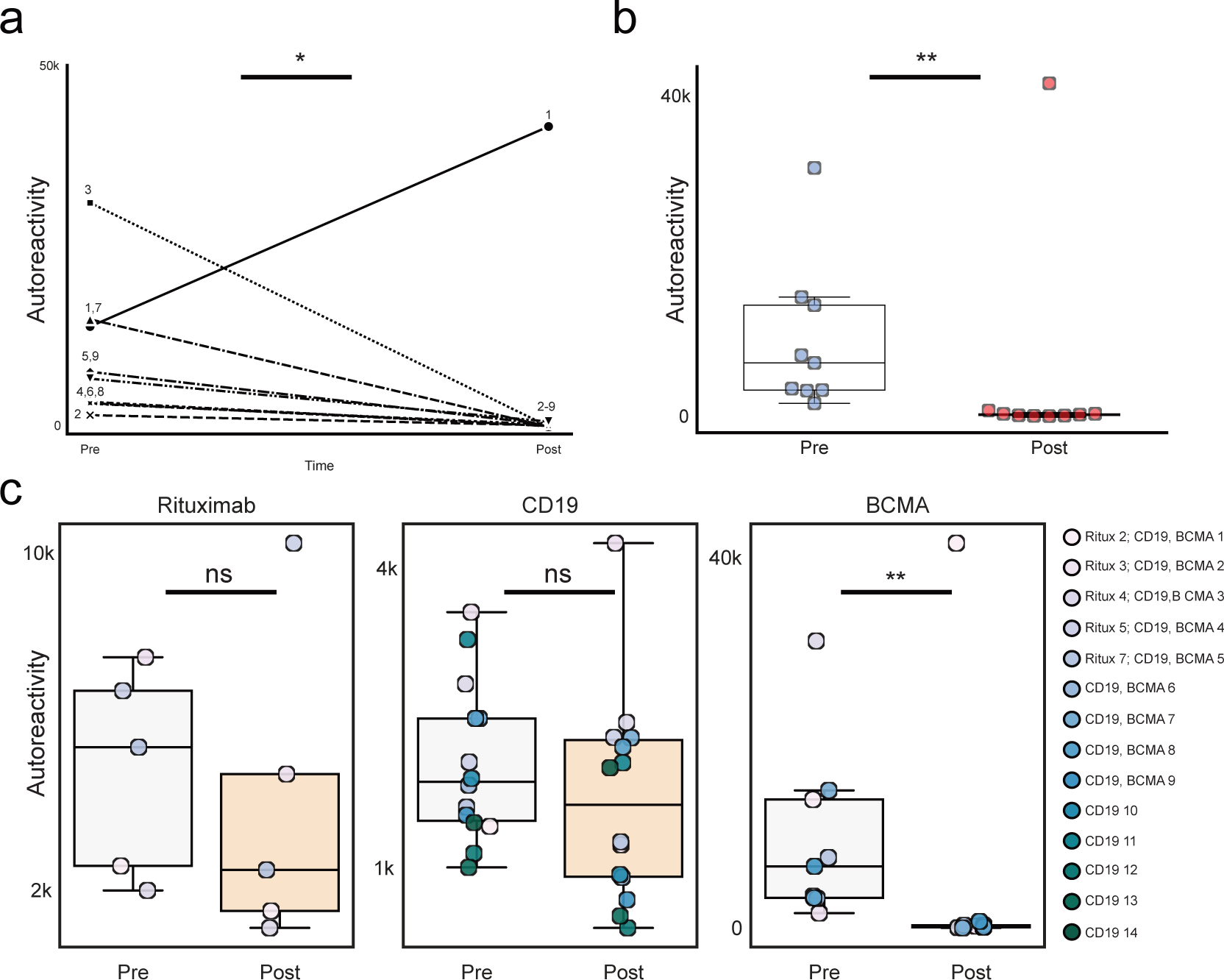
Changes in PhIP-Seq detected autoreactivities following anti-BCMA CAR-T cell therapy without controlling for potential confounding by paraprotein. **(a)** Lineplots showing the autoreactivity (sum of top 10 PhIP-Seq Z-scores relative to the 79 healthy controls) for each patient before and after treatment. One-sided paired samples Wilcoxon test p-value=0.049. **(b)** Swarmplots showing the relative distributions of autoreactivity (sum of top 10 PhIP-Seq Z-scores relative to the 79 healthy controls) before and after treatment with anti-BCMA CAR-T. Mann-Whitney U p-value=0.006. **(c)** Boxplots showing the relative distributions of autoreactivity (sum of top 10 PhIP-Seq Z-scores relative to the 79 healthy controls) before and after treatment with rituximab, anti-CD19 CAR-T, and anti-BCMA CAR-T. rituximab treatment cohort Mann-Whitney U p-value=0.42 with a median percent decrease of 52.3%; anti-CD19 CAR-T treatment cohort Mann-Whitney U p-value=0.206 with a median percent decrease of 11.9%; anti-BCMA CAR-T treatment cohort Mann-Whitney U p-value=0.006 with a median percent decrease of 97.2%.

**Extended Data Figure 7:**
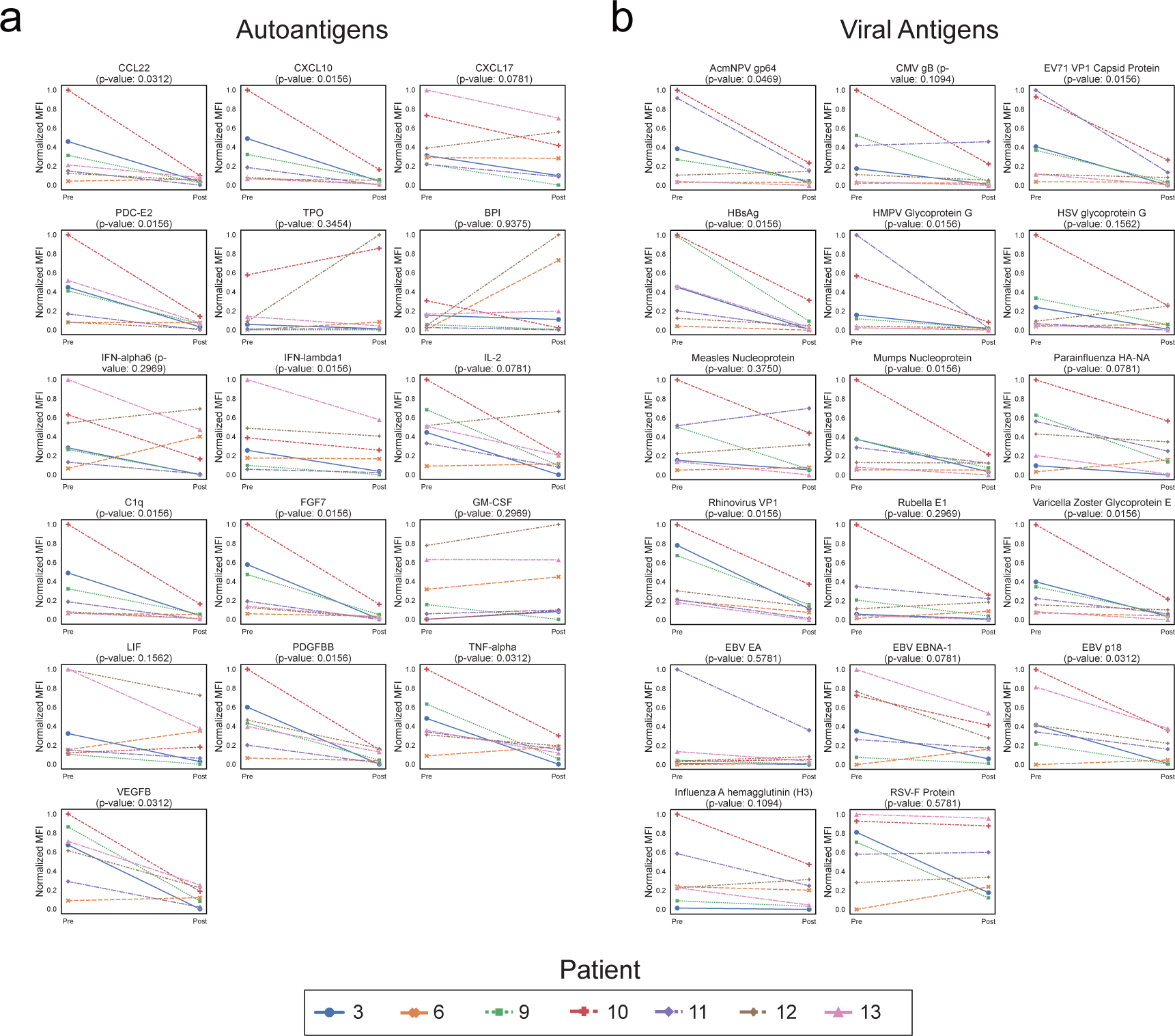
Changes in multiplex bead-based protein array detected antibody levels following BCMA therapy. **(a)** Multiplex bead-based protein array detected normalized MFI values (see “Methods”) for each of the 16 autoantigens with meaningful signal (see “Methods”) in each of the patients before and after treatment with anti-BCMA CAR-T cells. P-values shown are from a paired-samples Wilcoxon test. **(b)** Multiplex bead-based protein array detected normalized MFI values (see “Methods”) for each of the 17 viral antigens with meaningful signal (see “Methods”) in each of the patients before and after treatment with anti-BCMA CAR-T cells. P-values shown are from a paired-samples Wilcoxon test.

**Extended Data Figure 8:**
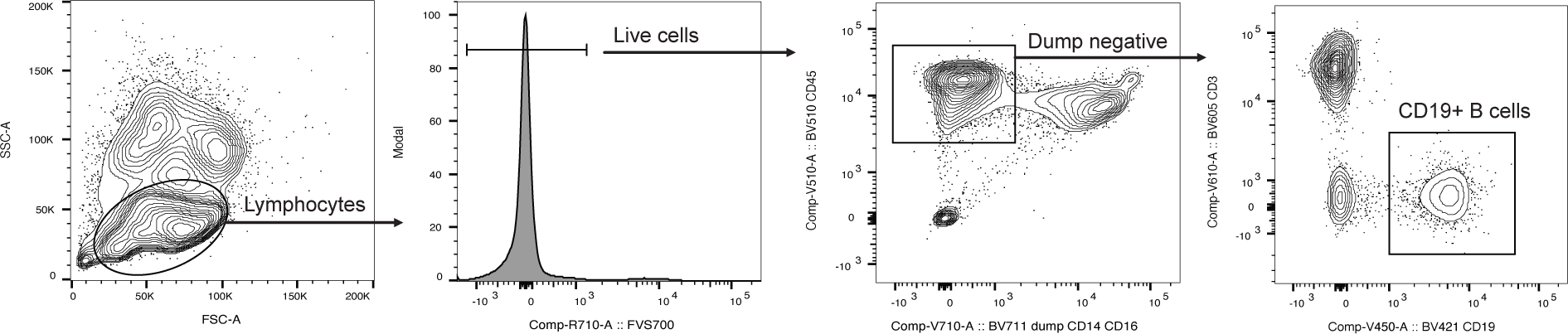
Flow cytometry gating strategy for identification of CD19^+^ B cells. Representative flow cytometry gating for the identification and enumeration of CD19^+^ B-cells.

**Extended Data Table 1:**
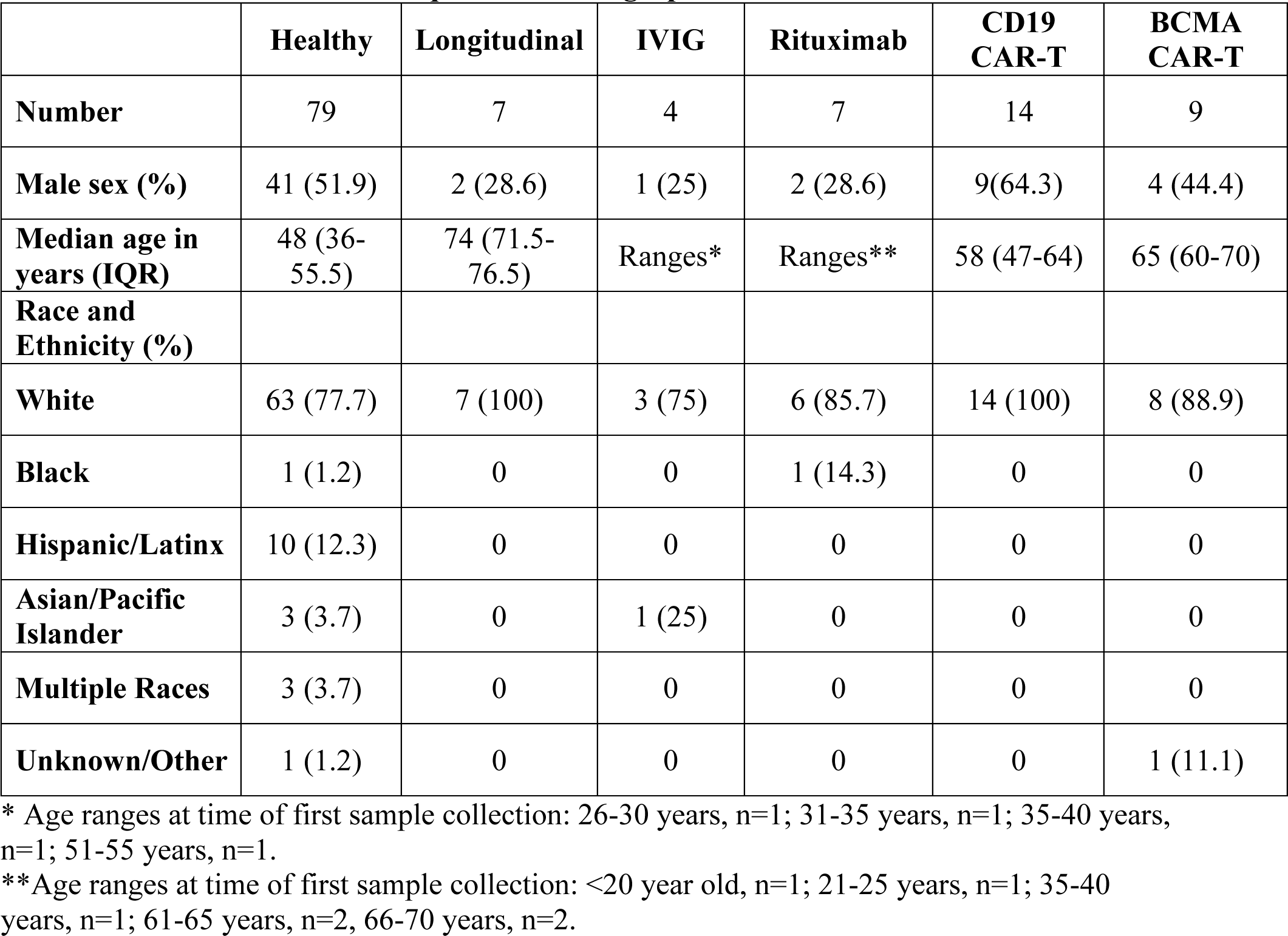
PhIP-Seq Cohort Demographics.

|  | Healthy | Longitudinal | IVIG | Rituximab | CD19 CAR-T | BCMA CAR-T |
| --- | --- | --- | --- | --- | --- | --- |
| Number | 79 | 7 | 4 | 7 | 14 | 9 |
| Male sex (%) | 41 (51.9) | 2 (28.6) | 1 (25) | 2 (28.6) | 9(64.3) | 4 (44.4) |
| Median age in years (IQR) | 48 (36-55.5) | 74 (71.5-76.5) | Ranges* | Ranges** | 58 (47-64) | 65 (60-70) |
| Race and Ethnicity (%) |  |  |  |  |  |  |
| White | 63 (77.7) | 7 (100) | 3 (75) | 6 (85.7) | 14 (100) | 8 (88.9) |
| Black | 1 (1.2) | 0 | 0 | 1 (14.3) | 0 | 0 |
| Hispanic/Latinx | 10 (12.3) | 0 | 0 | 0 | 0 | 0 |
| Asian/Pacific Islander | 3 (3.7) | 0 | 1 (25) | 0 | 0 | 0 |
| Multiple Races | 3 (3.7) | 0 | 0 | 0 | 0 | 0 |
| Unknown/Other | 1 (1.2) | 0 | 0 | 0 | 0 | 1 (11.1) |

\* Age ranges at time of first sample collection: 26-30 years, n=1; 31-35 years, n=1; 35-40 years, n=1; 51-55 years, n=1.
\*\*Age ranges at time of first sample collection: <20 year old, n=1; 21-25 years, n=1; 35-40 years, n=1; 61-65 years, n=2, 66-70 years, n=2.

**Extended Data Table 2:**
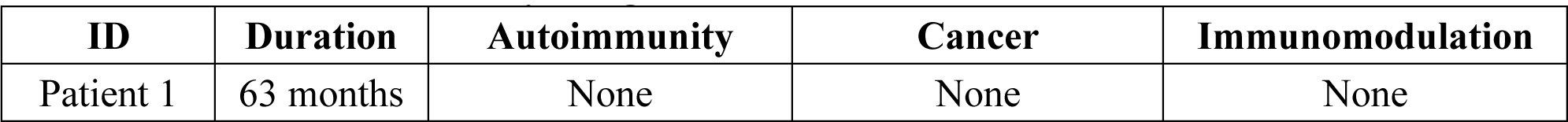
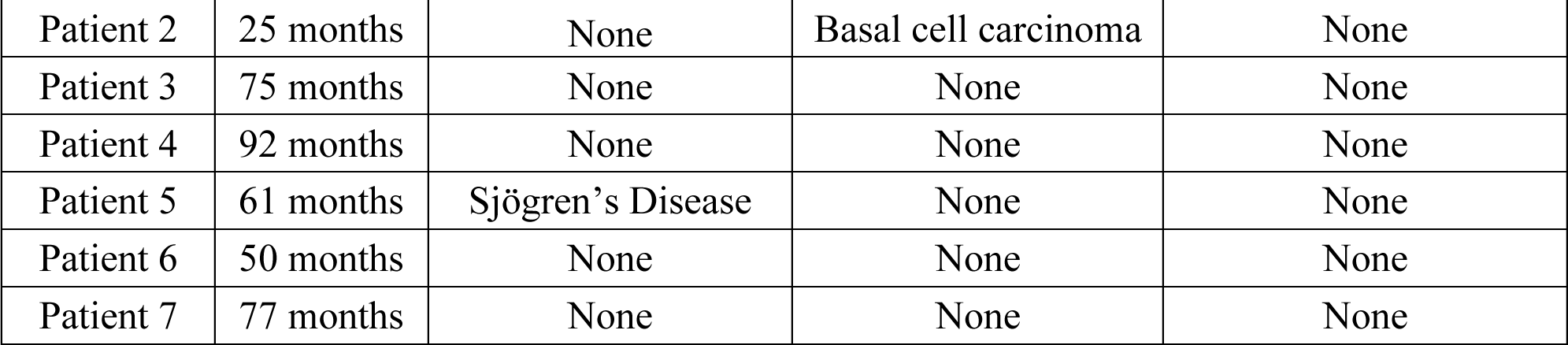
Healthy Longitudinal Patient Clinical Details.

**Extended Data Table 3:** IVIG Sample Details.

| ID | Sample Timing | Immunomodulation | Last IVIG Dose |
| --- | --- | --- | --- |
| **Patient 1** |  |  |  |
|  | Initial | None | Naïve |
|  | 6 weeks later | Prednisone/Imuran | 3 weeks prior |
| **Patient 2** |  |  |  |
|  | Initial | None | Naïve |
|  | 14 weeks later | Prednisone | 1 day prior |
| **Patient 3** |  |  |  |
|  | Initial | Prednisone | Naïve |
|  | 7 weeks later | Prednisone | 2 days prior |
| **Patient 4** |  |  |  |
|  | Initial | None | Naïve |
|  | 21 weeks later | Prednisone | 5 weeks prior |

**Extended Data Table 4:** Rituximab Sample Details.

| ID | Autoantibody | Sample | Immunomodulation | Rituximab |
| --- | --- | --- | --- | --- |
| **Patient 1** | **AchR** |  |  |  |
|  |  | 1 | Prednisone | Naïve |
|  |  | 2 | Prednisone | 1.1 |
|  |  | 3 | Prednisone | 3.9 |
|  |  | 4 | Prednisone | 3.4 |
|  |  | 5 | Prednisone | 6.1 |
|  |  | 6 | Prednisone | 24.6 |
| **Patient 2** | **MuSK** |  |  |  |
|  |  | 1 | Prednisone | Naïve |
|  |  | 2 | Prednisone | 2.3 |
|  |  | 3 | Prednisone | 5.1 |
|  |  | 4 | Prednisone | 4.8 |
| **Patient 3** | **MuSK** |  |  |  |
|  |  | 1 | Prednisone | 6.1 |
|  |  | 2 | Prednisone | 5.1 |
|  |  | 3 | None | 68.9 |
| **Patient 4** | **MuSK** |  |  |  |
|  |  | 1 | None | 8 |
|  |  | 2 | None | 7.9 |
|  |  | 3 | None | 24.7 |
| **Patient 5** | **MuSK** |  |  |  |
|  |  | 1 | None | 17.8 |
|  |  | 2 | None | 1.3 |
|  |  | 3 | None | 8.6 |
|  |  | 4 | None | 13.9 |
|  |  | 5 | None | 18.8 |
|  |  | 6 | None | 21 |
|  |  | 7 | None | 5.7 |
|  |  | 8 | None | 1.6 |
| **Patient 6** | **MuSK** |  |  |  |
|  |  | 1 | Prednisone | Naïve |
|  |  | 2 | Prednisone, Imuran | 1.7 |
|  |  | 3 | Prednisone, Imuran | 4.6 |
|  |  | 4 | Prednisone, Imuran | 4.7 |
|  |  | 5 | Prednisone, Imuran | 8.5 |
| **Patient 7** | **AChR** |  |  |  |
|  |  | 1 | Prednisone, Imuran | 5.7 |
|  |  | 2 | Prednisone, Imuran | 7 |
|  |  | 3 | None | 24.1 |

**Extended Data Table 5.** CAR-T Sample Details.

| **CAR-T** | **Patient** | **Malignancy** | **Post-CAR-T**<br>**Sample Timing**<br>**(weeks)** | **Prior therapy**<br>**(months pre-CAR-**<br>**T)** | **Concurrent**<br>**treatment (days**<br>**prior to second**<br>**sample)** |
| --- | --- | --- | --- | --- | --- |
| **BCMA** |  |  |  |  |  |
|  | Patient 1 | MM | 51 | IVIG, 6 | IVIG, 117 |
|  | Patient 2 | MM | 27 | IVIG, 4 | IVIG, 56 |
|  | Patient 3 | MM | 30 | None | None |
|  | Patient 4 | MM | 61 | None | IVIG, 68 |
|  | Patient 5 | MM | 29 | BCMA, 3 | IVIG, 112 |
|  | Patient 6 | MM | 29 | Daratumumab, 3 | IVIG, 117 |
|  | Patient 7 | MM | 30 | Daratumumab, 5 | IVIG, 47 |
|  | Patient 8 | MM | 22 | None | None |
|  | Patient 9 | MM | 22 | None | None |
| **CD19** |  |  |  |  |  |
|  | Patient 1 | Lymphoma | 33 | Polatuzumab, 1 | None |
|  | Patient 2 | Lymphoma | 28 | Autologous HCT, 7 | None |
|  | Patient 3 | Lymphoma | 32 | None | None |
|  | Patient 4 | Lymphoma | 36 | None | None |
|  | Patient 5 | Lymphoma | 27 | Ibrutinib, 1 | IVIG, 121 |
|  | Patient 6 | Lymphoma | 30 | None | None |
|  | Patient 7 | Lymphoma | 29 | None | IVIG, 59 |
|  | Patient 8 | Lymphoma | 26 | None | None |
|  | Patient 9 | Lymphoma | 30 | None | IVIG, 28 |
|  | Patient 10 | Lymphoma | 35 | Autologous HCT, 8 | None |
|  | Patient 11 | Lymphoma | 30 | None | IVIG, 28 |
|  | Patient 12 | Lymphoma | 21 | None | None |
|  | Patient 13 | Lymphoma | 27 | None | None |
|  | Patient 14 | Lymphoma | 27 | None | IVIG, 155 |

**Table Legend:** MM=multiple myeloma; Sample timing refers to number of weeks between CAR-T infusion and the post-infusion sample being drawn; Prior therapy lists additional immunomodulatory therapies given in the year prior to initial sample collection, and the number of months between administration of the therapy and collection of the initial sample, excluding the pre-CAR-T conditioning chemotherapy which consists of cyclophosphamide and fludarabine for every patient; Concurrent treatment lists the number of days prior to the second sample being collected when treatment was administered, only IVIG is listed because no other B cell or antibody modulating therapies were given.

**Extended Data Table 6:**
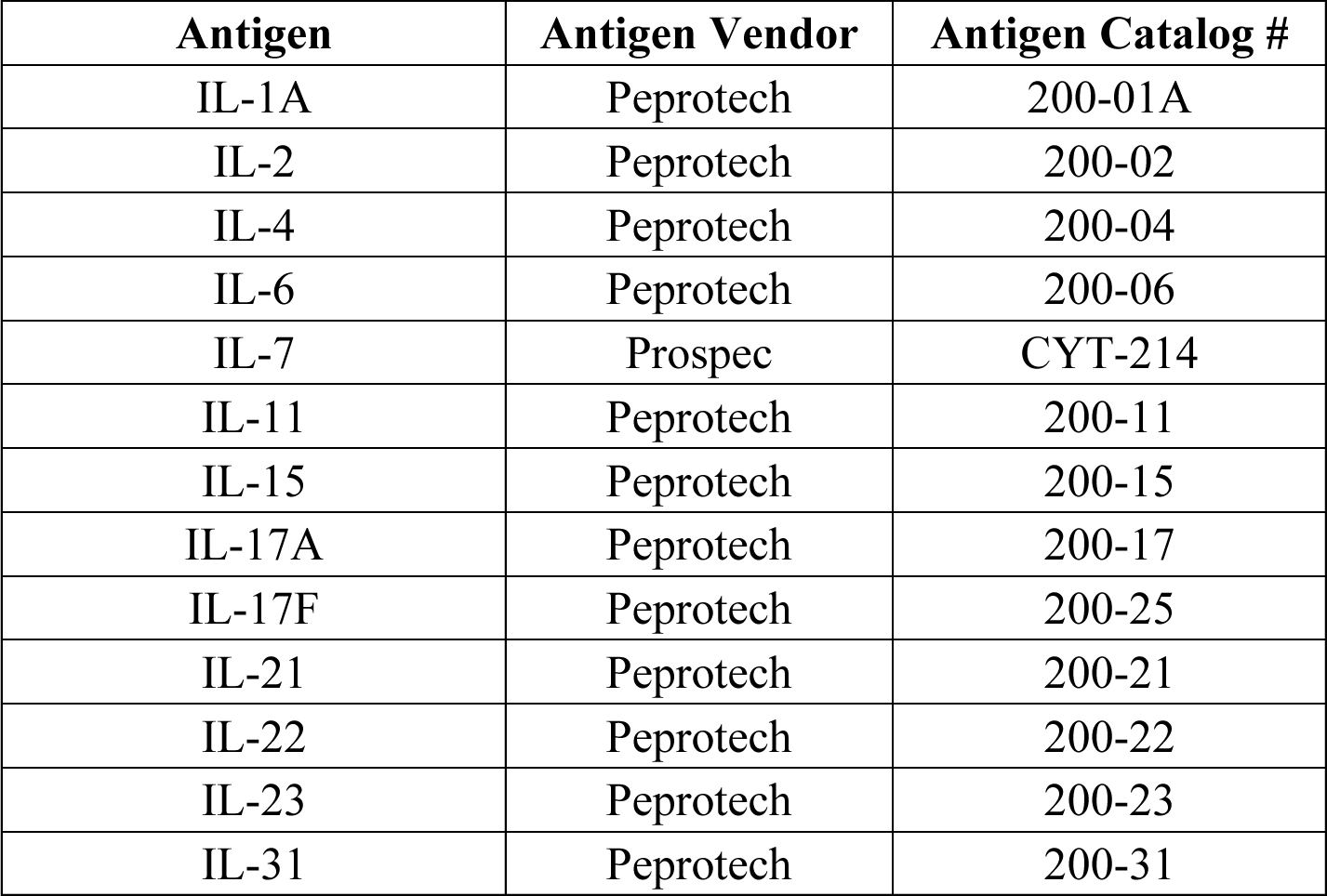
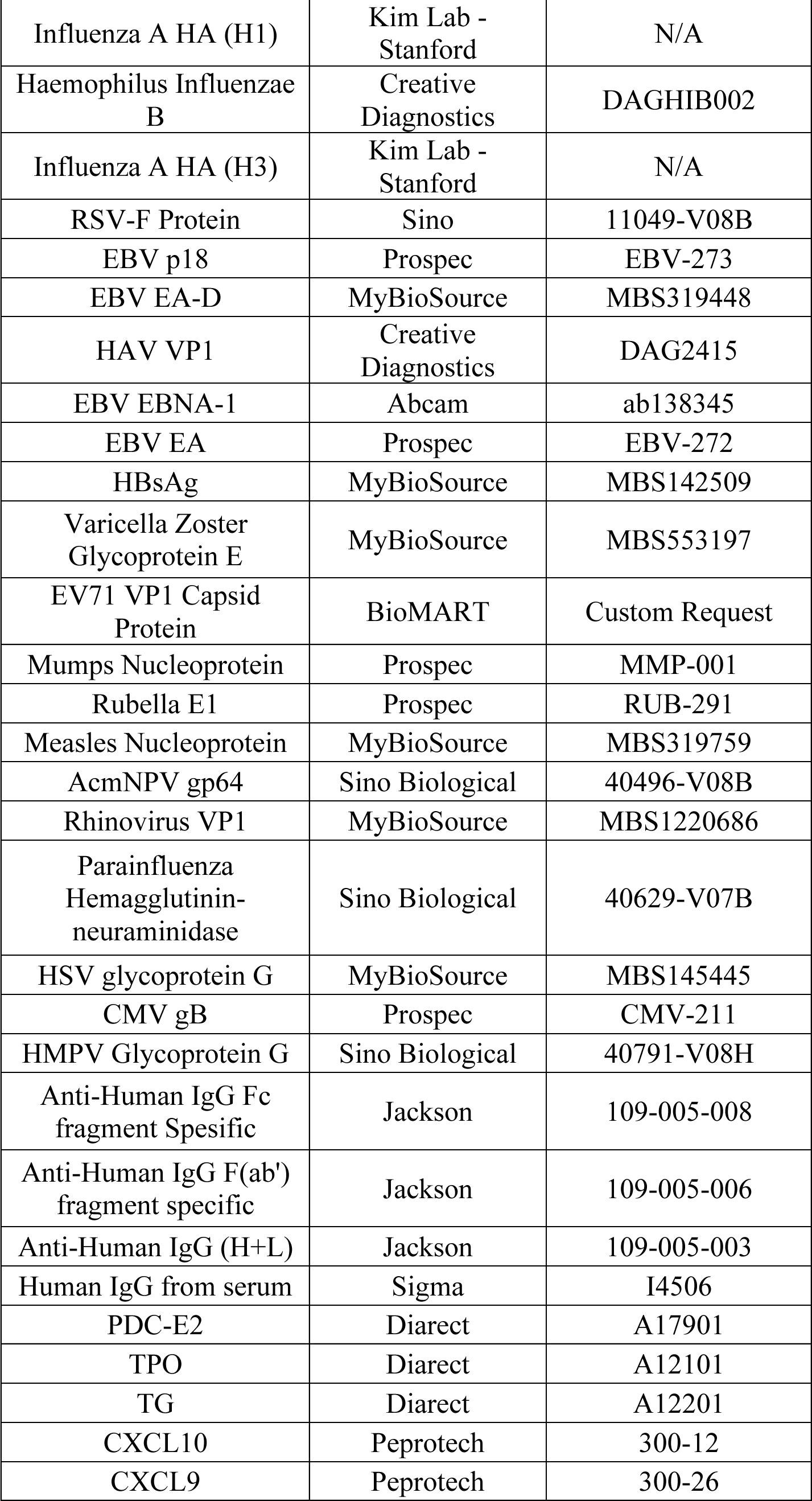
Multiplex Bead-based Protein Array Antigen Panel.

**Extended Data Table 7:**
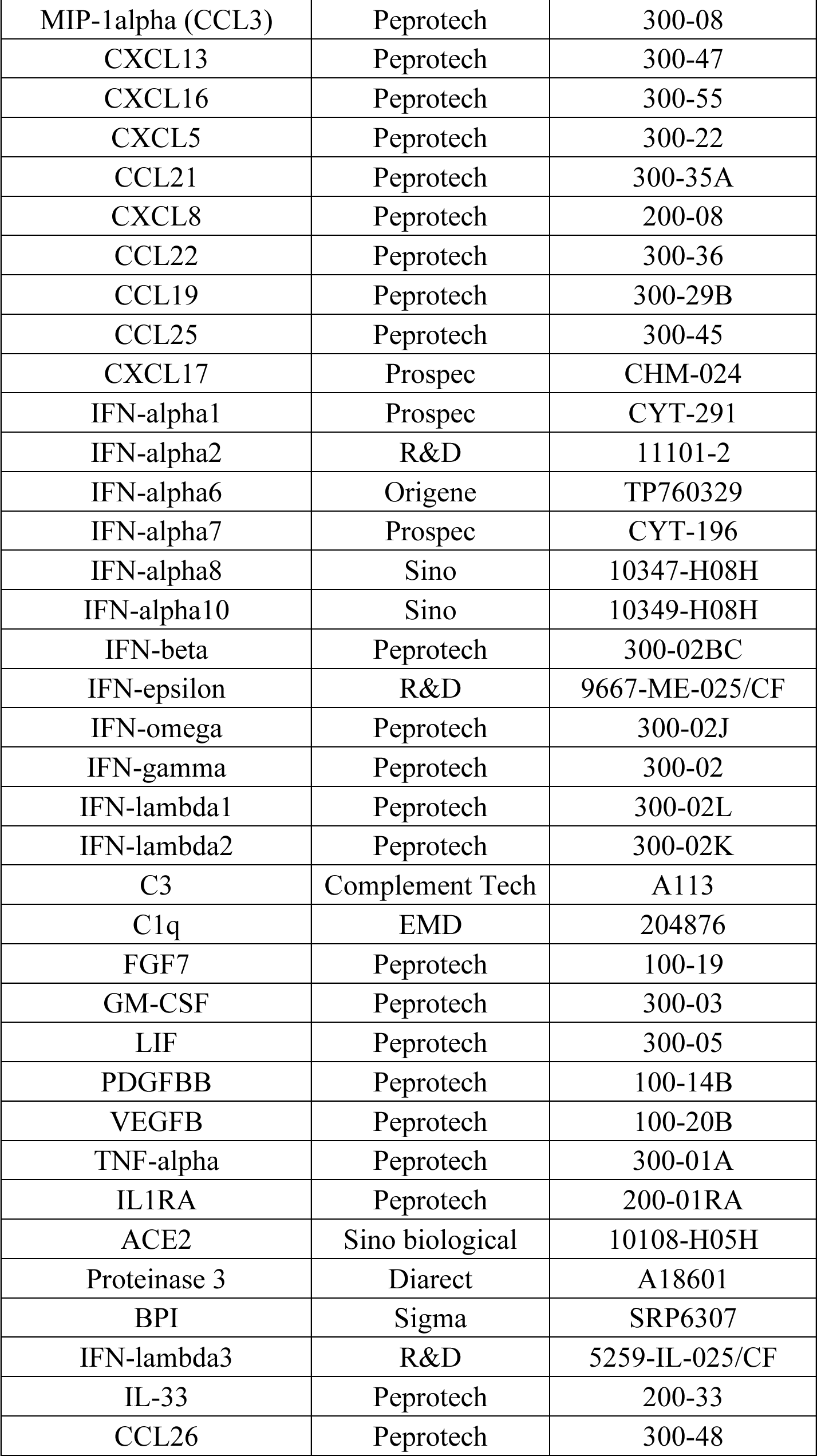
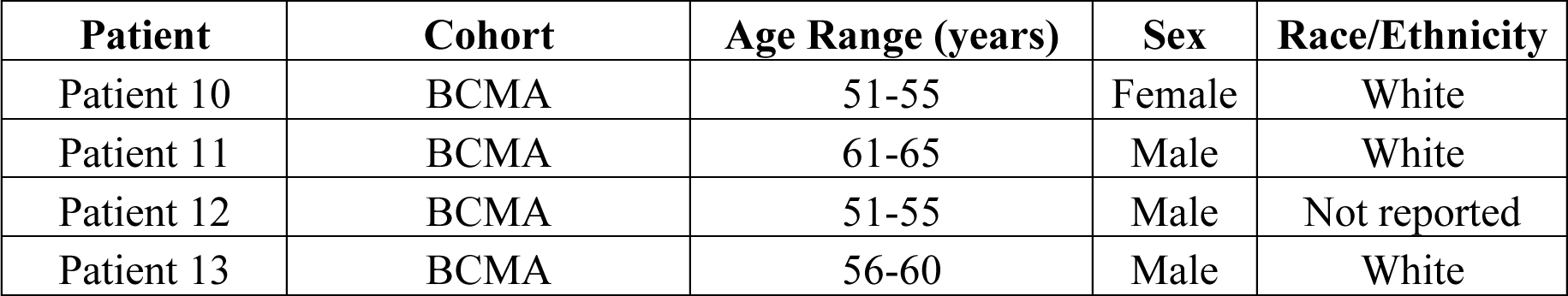
Additional BCMA Patient Demographics.

**Extended Data Table 8:** Additional BCMA Sample Details.

| CAR-T | Patient | Malignancy | Post-CAR-T<br>Sample Timing<br>(weeks) | Prior therapy<br>(months pre-CAR-T) | Concurrent<br>treatment (days<br>prior to second<br>sample) |
| --- | --- | --- | --- | --- | --- |
| BCMA |  |  |  |  |  |
|  | Patient 10 | MM | 27 | None | None |
|  | Patient 11 | MM | 24 | None | None |
|  | Patient 12 | MM | 27 | None | None |
|  | Patient 13 | MM | 22 | Elotuzumab, 4 | None |

**Table Legend:** MM=multiple myeloma; Sample timing refers to number of weeks between CAR-T infusion and the post-infusion sample being drawn; Prior therapy lists additional immunomodulatory therapies given in the year prior to initial sample collection, and the number of months between administration of the therapy and collection of the initial sample, excluding the pre-CAR-T conditioning chemotherapy which consists of cyclophosphamide and fludarabine for every patient.

## Notes

### Author Declarations

Ethics committee/IRB (IRB #10-02076) of University of California in San Francisco gave ethical approval for this work. Ethics committee/IRB (Protocol 10080) of the Fred Hutchinson Cancer Center gave ethical approval for this work. Ethics committee/IRB (EXPLORE-MG Registry, NCT03792659) of Yale University gave ethical approval for this work.

